# Effectiveness of telehealth low-carbohydrate intervention in preventing chronic kidney disease: a real-world, retrospective, matched cohort study

**DOI:** 10.1101/2025.10.17.25338238

**Authors:** Shaminie J Athinarayanan, Petter Bjornstad, Priya V Shanmugam, Thomas Weimbs, Adam J Wolfberg, Jeff S Volek, Jonathan Himmelfarb, Richard J Johnson

**Affiliations:** Research Department, Virta Health, Denver, CO; Kidney Research Institute and Division of Metabolism, Endocrinology, and Nutrition (need to change this in the proof), Department of Medicine, University of Washington, Seattle, WA, USA; Department of Molecular, Cellular, and Developmental Biology, University of California, Santa Barbara, CA, USA; Department of Human Sciences, College of Education and Human Ecology, The Ohio State University, Columbus, OH, USA; Department of Medicine, Icahn School of Medicine at Mount Sinai, New York, NY, USA; Division of Renal Diseases and Hypertension, Department of Medicine, University of Colorado Anschutz Medical Campus, Aurora, CO, USA

**Keywords:** Chronic kidney disease, telemedicine, carbohydrate reduction, ketosis, type 2 diabetes, obesity

## Abstract

**Background:** Lifestyle interventions targeting metabolic health may influence chronic kidney disease (CKD) risk, particularly among adults with type 2 diabetes and obesity. We evaluated real-world associations between a telehealth-delivered, individualized nutrition therapy program (VINT) and CKD incidence and progression compared with usual care.

**Methods:** We conducted a retrospective, propensity score–matched cohort study using the Komodo Healthcare Map™, a longitudinal U.S. administrative claims database. Adults enrolled in a telehealth-delivered nutrition therapy emphasizing carbohydrate reduction, VINT (n=11,077) were matched 1:1 to usual-care controls on demographic, clinical, and medication covariates, over five years of follow-up. Primary outcomes were new-onset CKD, CKD stage ≥3, and CKD stage ≥4. Secondary outcomes included renal safety events. Cox proportional hazards and Poisson regression models were used to estimate hazard ratios (HRs) and incidence rate ratios (IRRs).

**Results:** VINT participation was associated with a lower incidence of new-onset CKD (10.1 vs 15.6 per 1,000 person-years; HR 0.64, 95% CI 0.53–0.77; p<0.001), CKD stage ≥3 (HR 0.57, 95% CI 0.45–0.73; p<0.001), and CKD stage ≥4 (HR 0.38, 95% CI 0.18–0.79; p=0.009). No increased risk of kidney stones, metabolic acidosis, diabetic ketoacidosis, or gout was observed in VINT compared with the UC group.

**Conclusions:** In this large, real-world matched cohort, participation in a telehealth-delivered nutrition therapy emphasizing carbohydrate reduction was associated with lower CKD incidence and progression without increased renal adverse events. These findings suggest that individualized nutrition therapy may be a safe and scalable approach to CKD prevention and management in adults with metabolic disease, warranting further prospective investigation.

## Introduction

The prevalence of chronic kidney disease (CKD) has risen sharply, particularly among individuals with type 2 diabetes (T2D) (1,2). CKD carries profound consequences, including reduced quality of life, elevated cardiovascular risk, and increased morbidity and mortality (2–4). Although recent therapeutic advances including sodium-glucose cotransporter 2 inhibitors (SGLT2i) (5,6), nonsteroidal mineralocorticoid receptor antagonists (7), and GLP1 receptor agonists (GLP1-RA) (8,9) offer benefit, medical therapy remains the foundation of standard care, and these agents are costly, may cause adverse effects, and leave substantial residual risk of CKD progression (1,2,10). Accordingly, lifestyle interventions, particularly dietary strategies, remain critical as complementary approaches either alone or in combination with pharmacotherapy.

High-protein diets accelerate CKD progression in animals (11,12), prompting investigators to study protein restriction as an intervention (13). However, benefits were modest or limited to subgroups, and extreme restriction was found to have deleterious effects on overall morbidity and mortality (13). Current guidelines therefore recommend avoiding high protein intake but emphasize only modest restriction (0.6-1.0 g/kg/day) for individuals with moderate CKD (14). In contrast, the 2020 KDOQI guidelines recommend more stringent protein restriction (as low as 0.55-0.60 g/kg/day in metabolically stable patients), highlighting ongoing uncertainty and debate regarding the optimal level of dietary protein intake in CKD (15). Similarly, caloric restriction may confer some benefit in CKD linked to metabolic syndrome or diabetes (16). Despite investigation, dietary therapy has not been established as a disease-modifying standard of care for CKD. Evidence, however, implicates high carbohydrate intake in CKD progression. Carbohydrate excess contributes to metabolic syndrome and diabetes, and experimental studies demonstrate that diets high in sugar or fructose can accelerate CKD (17,18).

We studied a remotely administered, digitally supported nutrition and lifestyle program developed primarily for individuals with type-2 diabetes and obesity/overweight that focuses on carbohydrate reduction leading to varying degrees of nutritional ketosis (19,20). This intervention is delivered in the context of usual medical care; however, adherence to the program is associated with a high rate of type 2 diabetes remission, often accompanied by complete medication deprescription (19,20). Post-hoc analyses of clinical trial data from this intervention also demonstrated a reversal of eGFR slope decline, with a significant increase in eGFR from baseline to two years, corresponding to a slope of +0.91 mL/min/1.73m²/year (21). The multimodal intervention, integrating personalized carbohydrate reduction with continuous remote care, individualized treatment, and real-time clinical support, likely achieved these effects by addressing multiple interrelated mechanisms rather than isolated pathways. Beyond eGFR benefits, both clinical trials and real-world studies show improvements in glycemia (19,20,22), weight (19,20,22), and blood pressure (23). Kidney-specific benefits include durable reductions in urinary albumin-to-creatinine ratio (UACR) among patients with stage 2–3 albuminuria, sustained for up to two years (24).

In this claims-based analysis, we evaluated associations between participation in a multimodal, carbohydrate-reduced lifestyle intervention delivered alongside usual care and the development and progression of CKD, compared with matched individuals receiving usual care alone. Given the limitations of claims data without linked laboratory data, this analysis was designed to be exploratory and hypothesis-generating, intended to estimate associations and effect sizes rather than establish causality. Outcomes included incidence of new CKD diagnoses, including albuminuria, and the occurrence of advanced CKD stages during follow-up. Safety outcomes included kidney stone, gout, and acidosis including diabetic ketoacidosis (DKA) diagnoses. Finally, we explored whether metabolic response, defined by ketone levels, weight loss, and A1c change, was associated with CKD risk within VINT cohort and stability or regression of existing CKD.

## Materials and Methods

### Data Source

This study used the Komodo Healthcare Map™, a longitudinal U.S. claims database covering ∼330 million individuals across Medicare, Medicaid, and commercial payers (25). Program participant data were linked to claims to enable longitudinal follow-up. A control cohort was drawn from the same database and aligned to the treated cohort on key characteristics. Analyses were conducted within the Komodo Sentinel environment (data version: April 19, 2026). Additional details are provided in the Supplementary Methods.

### Study Population and Design

The study included adults enrolled in the Virta Individualized Nutrition Therapy (VINT) program with type 2 diabetes, overweight, or obesity. VINT consisted of continuous remote care integrating individualized nutrition therapy, medication management, and health coaching. Each participant in VINT works with a health coach who monitors dietary adherence and supports behavioral change, while an assigned clinician oversees medical management, including addressing clinical concerns and adjusting medications, as needed. Further details are provided in the Supplementary Methods.

Eligible participants in both VINT and usual care (UC) cohorts were required to have ≥1 year of continuous pre-index claims coverage and ≥6 months of claims follow-up. Sequential exclusions removed individuals with conditions incompatible with the VINT or likely to confound CKD outcomes (e.g., type 1 diabetes, severe comorbid illness, or non-CKD kidney diseases). Those with any baseline CKD and albuminuria were excluded as well.

The control cohort was constructed by assigning index dates aligned to the treated cohort’s enrollment distribution and applying identical inclusion and exclusion criteria. UC control was matched 1:1 to VINT participants using propensity scores based on demographic, clinical, and healthcare utilization variables. The primary analysis cohort included all eligible VINT participants with available closed claims data and their matched UC controls, regardless of program duration, to enhance generalizability. A secondary cohort was constructed by restricting the analysis to participants with ≥6 months of continuous VINT participation and rematching UC controls using the same matching approach as the primary analysis. The secondary cohort was used for exploratory analyses to evaluate outcomes among participants with sufficient exposure to VINT. Outcomes were assessed from the index date in both cohorts. Full eligibility criteria and matching specifications are detailed in the Supplementary Methods.

### Intervention and Medication Management

VINT delivered continuous remote care via telemedicine, combining personalized carbohydrate reduction, medication management, and ongoing health coaching, with regular monitoring of biomarkers to guide individualized adjustments.

Medications were actively managed by clinicians, with early reductions in hypoglycemia-prone drugs and ongoing adjustments based on patient response and real-time data. Comprehensive details on VINT and medication management are provided in the Supplementary Methods.

### Baseline Characteristics

Baseline variables included demographics, comorbidities, medication use, and healthcare utilization, defined using administrative claims (ICD-10-CM and drug codes). These variables were selected a priori and incorporated into the matching process. Detailed descriptions and definitions are provided in Supplementary Tables S1–S3 and Supplementary method.

### Study Outcomes

The primary outcomes were time to first occurrence of: (1) incident chronic kidney disease (CKD), defined as a new diagnosis of CKD stage 1–5, end-stage kidney disease (ESKD), renal dialysis, or albuminuria among participants without CKD or albuminuria at baseline; and (2) incident CKD stage 3 or higher, defined as a first diagnosis of CKD stage 3a, stage 3b, stage 4, stage 5, ESKD, or renal dialysis during follow-up. An exploratory outcome was incident CKD stage 4 or higher, defined as a first diagnosis of CKD stage 4, stage 5, ESKD, or renal dialysis during follow-up. Participants were censored at loss of continuous claims coverage or study end.

Stage-specific outcomes were defined to capture progression to more advanced CKD stages; individuals developing lower stages during follow-up were censored at the time lower stage occurred and not counted as events.

The secondary outcomes were the occurrence of kidney stones, acidosis including diabetic ketoacidosis, and gout during follow-up. Exploratory outcomes included: (1) assessment of predictors associated with new-onset CKD diagnosis in the VINT group from the secondary matched cohort, and (2) to evaluate CKD stage trajectories, we constructed a separate matched cohort of participants with baseline CKD and/or albuminuria. Among those with at least one follow-up CKD diagnosis prior to censoring, we assessed CKD stage transitions, including regression, stability, and progression, in both the VINT and UC groups. Detailed ICD-10-CM diagnosis codes used to define CKD outcomes are provided in Supplementary Table S1. Additional ICD-10-PCS (International Classification of Diseases, Tenth Revision, Procedure Coding System), CPT (Current Procedural Terminology), and HCPCS (Healthcare Common Procedure Coding System) procedure codes used to identify different exclusion criteria and advanced kidney disease ascertainment are provided in Supplementary Table S2.

### Statistical analysis

A full description of the statistical methods is provided in the Supplementary Materials and Methods. The study design, conduct, and reporting adhered to STROBE guidelines.

Categorical variables were summarized using counts and percentages, and continuous variables using means and standard deviations. UC controls were matched 1:1 to VINT participants using nearest-neighbor propensity score matching without replacement based on demographic, clinical, medication, and healthcare utilization variables, with other comorbidities (e.g., type 2 diabetes, obesity, index year, relevant medications, etc) enforced as exact matches. Additional details regarding the matching procedure and balance assessment are provided in the Supplementary Methods. Outcomes were assessed from the index date.

For primary outcomes, Cox proportional hazards regression was used to estimate hazard ratios (HRs) with 95% confidence intervals (CIs). Models were adjusted for age, sex, race/ethnicity, and baseline comorbidities. Different modeling strategy was applied to assess robustness: (1) a base model without medication adjustment; (2) models additionally adjusting for follow-up medication use (modeled as ever/never and proportion of days covered); and (3) models incorporating medication use as a time-varying covariate. Additional sensitivity analyses using marginal structural models with inverse probability weighting were conducted to account for time-varying confounding and potential mediation by medication use. Outcomes were assessed in both the primary and secondary cohorts, as defined in the study outcomes. As a further robustness check, inverse probability of treatment weighting (IPTW) was applied, retaining all eligible VINT participants and UC. Given the potential for mortality to act as a competing event for CKD outcomes, we also evaluated all incident CKD outcomes using Fine–Gray subdistribution hazard models. In addition, mortality was assessed as a separate outcome within the study cohort. Incidence rates were also calculated as the number of events per 1,000 person-years of follow-up. Person-years were based on total observed follow-up time from the index date and were used for descriptive purposes only. Additional details are provided in the Supplementary Methods.

Analyses of secondary safety outcomes and exploratory endpoints, including adverse events, predictors of CKD, and CKD stage transitions, are summarized in the Supplementary Methods. Statistical significance was defined as a two-sided α of 0.05. All analyses were conducted using R.

## Results

### Study Participant Flow

A total of 115,942 VINT participants and 2.39 million controls were identified in the Komodo Sentinel database (Figure 1; Supplementary Table S4). After applying inclusion criteria, 11,890 VINT and 310,889 UC participants remained. Following exclusions and after removing those with baseline CKD or albuminuria diagnosis, 11,255 VINT participants (after removing duplicates) and 274,455 UC controls were eligible for propensity score matching.

**Figure 1.**
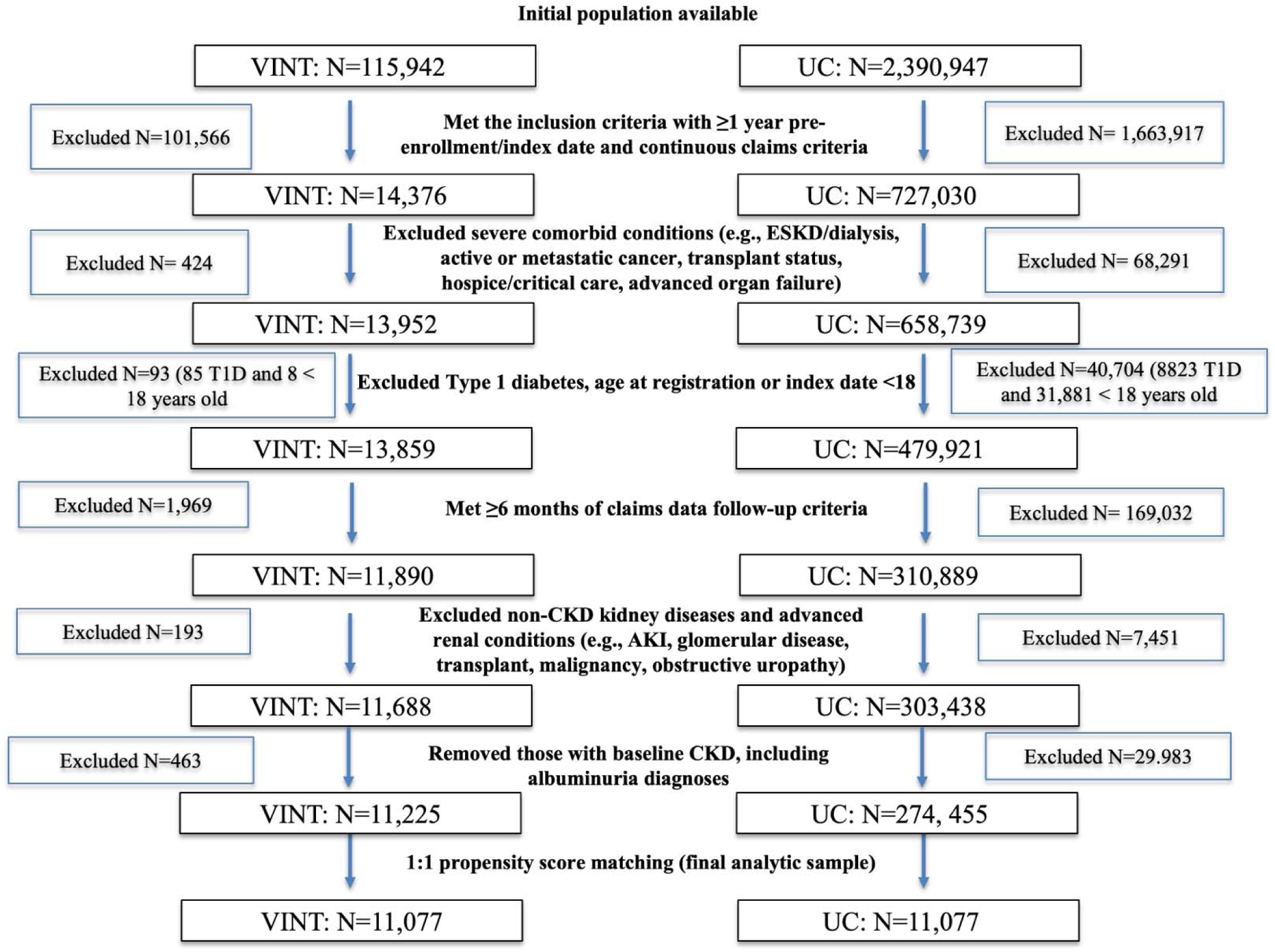
Patient attrition flowchart from the initial population in the Komodo Healthcare Sentinel working environment to the final analytic sample after 1:1 matching.

Using a stringent (Tier 3) 1:1 propensity score–matching approach, the final analytic sample included 11,077 VINT participants and 11,077 matched UC.

### Baseline Characteristics

Baseline characteristics were well balanced between groups in the primary matched cohort, with all standardized mean differences (SMDs) <0.1 (Tables 1 and 2). The mean age was 51 years, with a female-to-male ratio of approximately 3:2. Mean follow-up was 1.8 years for VINT and 1.9 years for UC. Control index dates were aligned with VINT enrollment years to account for temporal trends (Supplementary Figure S1). The matched VINT cohort was broadly representative of excluded VINT participants (Supplementary Table S5), with baseline characteristics largely balanced (SMDs <0.1), except for minor differences in race/ethnicity and enrollment index year, which were addressed in matching. Similarly, baseline characteristics in the secondary matched cohort were also well balanced between groups, with all SMDs <0.1 (Supplementary Table S6).

**Table 1.**
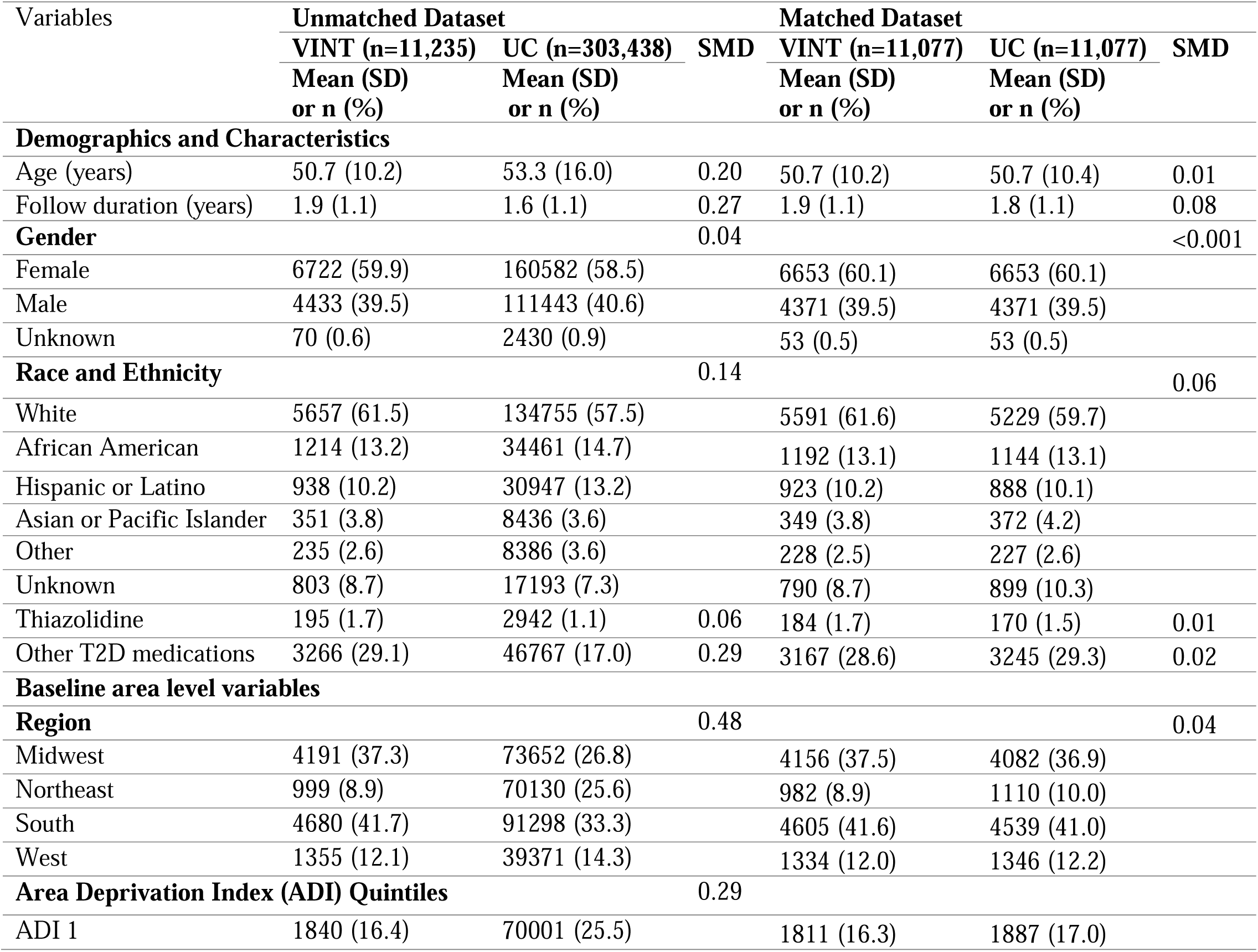
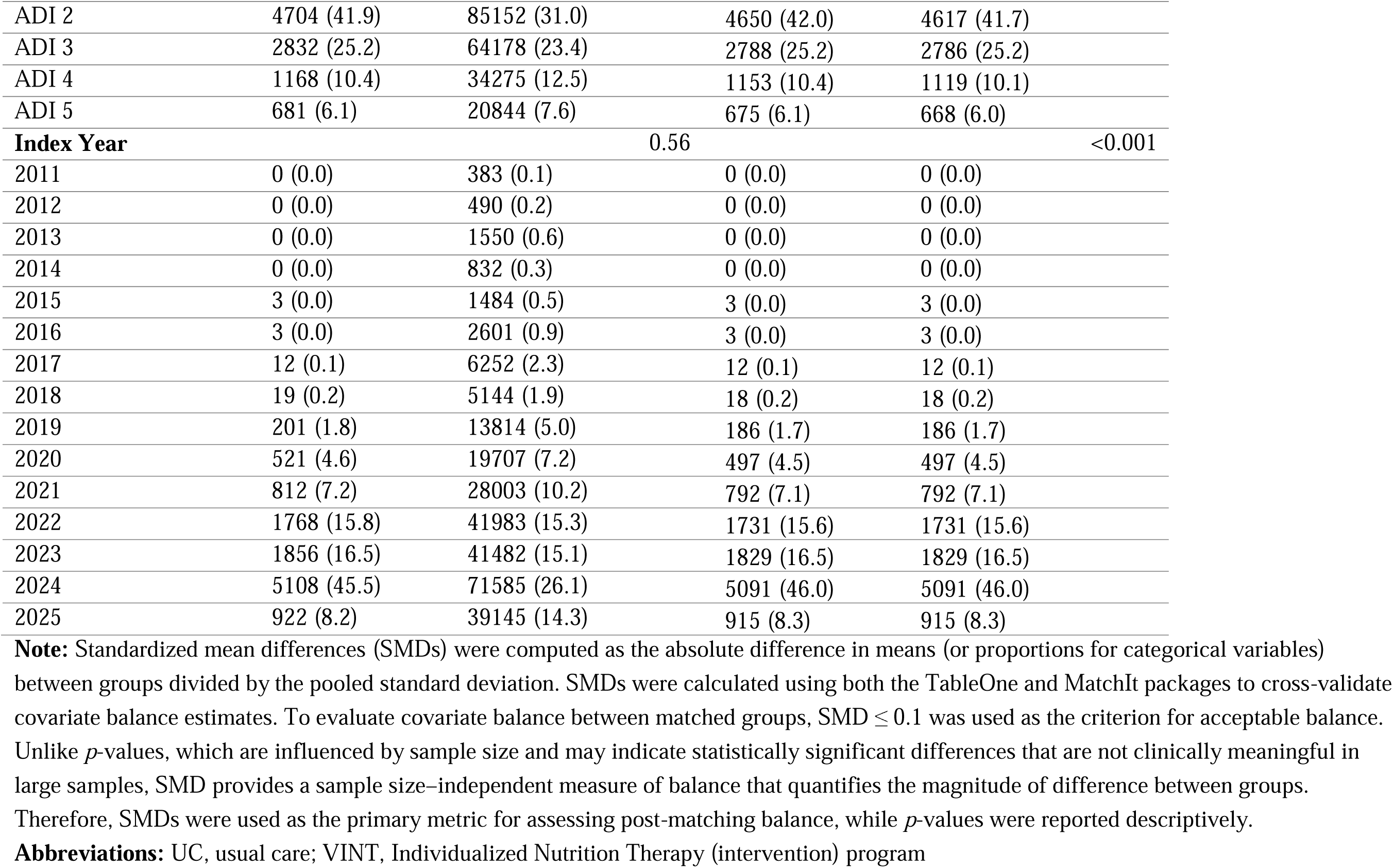
Baseline demographics, disease trajectory as assessed by cost, and area level variables of propensity score 1:1 matched cohort of VINT and UC.

**Table 2.** Baseline comorbidities and medication use of propensity score 1:1 matched cohort of VINT and UC, including those not used in the matching.

| Variables | Unmatched Dataset |  |  | Matched Dataset |  |  |
| --- | --- | --- | --- | --- | --- | --- |
|  | VINT (n=11,235) | UC (n=274,455) | SMD | VINT (n=11,077) | UC (n=11,077) | SMD |
|  | Mean (SD)<br>or n (%) | Mean (SD)<br>or n (%) |  | Mean (SD)<br>or n (%) | Mean (SD)<br>or n (%) |  |
| Baseline Comorbidities |  |  |  |  |  |  |
| Type 2 diabetes | 4086 (36.4) | 63283 (23.1) | 0.30 | 3976 (35.9) | 3976 (35.9) | <0.001 |
| Obesity | 6135 (54.7) | 125775 (45.8) | 0.18 | 6082 (54.9) | 6082 (54.9) | <0.001 |
| Chronic Kidney Disease | 0 (0.0) | 0 (0.0) | <0.001 | 0 (0.0) | 0 (0.0) | <0.001 |
| Kidney Stones | 179 (1.6) | 5525 (2.0) | 0.03 | 133 (1.2) | 133 (1.2) | <0.001 |
| Heart Failure | 87 (0.8) | 8874 (3.2) | 0.18 | 83 (0.7) | 91 (0.8) | 0.01 |
| Hyperlipidemia | 4957 (44.2) | 126180 (46.0) | 0.04 | 4868 (43.9) | 4978 (44.9) | 0.02 |
| Hypertension | 4757 (42.4) | 126826 (46.2) | 0.08 | 4671 (42.2) | 4637 (41.9) | 0.01 |
| Stroke | 153 (1.4) | 10059 (3.7) | 0.15 | 147 (1.3) | 139 (1.3) | 0.01 |
| Coronary artery disease | 484 (4.3) | 23203 (8.5) | 0.17 | 475 (4.3) | 420 (3.8) | 0.03 |
| Cardiovascular disease | 66 (0.6) | 3968 (1.4) | 0.09 | 65 (0.6) | 65 (0.6) | <0.001 |
| Smoking | 364 (3.2) | 17622 (6.4) | 0.15 | 353 (3.2) | 420 (3.8) | 0.03 |
| Baseline Medications |  |  |  |  |  |  |
| SGLT2i | 908 (8.1) | 11301 (4.1) | 0.17 | 842 (7.6) | 842 (7.6) | <0.001 |
| Incretin Mimetics | 1993 (17.8) | 31395 (11.4) | 0.18 | 1908 (17.2) | 1908 (17.2) | <0.001 |
| RAASi | 3701 (33.0) | 92801 (33.8) | 0.02 | 3620 (32.7) | 3620 (32.7) | <0.001 |
| MRA and nonMRA | 289 (2.6) | 7960 (2.9) | 0.02 | 241 (2.2) | 241 (2.2) | <0.001 |
| Diuretic | 2014 (17.9) | 60152 (21.9) | 0.10 | 1961 (17.7) | 1998 (18.0) | 0.01 |
| Beta blocker | 1224 (10.9) | 47955 (17.5) | 0.19 | 1192 (10.8) | 1164 (10.5) | 0.01 |
| Calcium blocker | 1282 (11.4) | 43893 (16.0) | 0.13 | 1248 (11.3) | 1175 (10.6) | 0.02 |
| Other antihypertensive medications | 230 (2.0) | 9051 (3.3) | 0.08 | 221 (2.0) | 209 (1.9) | 0.01 |
| Insulin | 605 (5.4) | 10047 (3.7) | 0.08 | 578 (5.2) | 673 (6.1) | 0.04 |
| Sulfonylureas | 339 (3.0) | 4901 (1.8) | 0.08 | 322 (2.9) | 344 (3.1) | 0.01 |
| DPP4i | 384 (3.4) | 5022 (1.8) | 0.10 | 368 (3.3) | 414 (3.7) | 0.02 |
| Thiazolidine | 195 (1.7) | 2942 (1.1) | 0.06 | 184 (1.7) | 170 (1.5) | 0.01 |
| Other T2D medications | 3266 (29.1) | 46767 (17.0) | 0.29 | 3167 (28.6) | 3245 (29.3) | 0.02 |
| **Baseline cost and trajectory** |  |  |  |  |  |  |
| Total cost of care | 11773.0 (24464.4) | 10649.2 (22926.6) | 0.05 | 11689.2 (24507.1) | 11603.19 (23553.7) | 0.004 |
| Outpatient cost | 904.5 (7410.2) | 1157.1 (7761.8) | 0.03 | 899.8 (7396.0) | 921.3 (6062.6) | 0.003 |
| Inpatient cost | 3638.8 (12464.6) | 3214.3 (12949.7) | 0.03 | 3557.3 (12448.5) | 3726.7 (11220.6) | 0.01 |
| Pharmacy cost | 7227.6 (17690.8) | 6271.3 (14965.2) | 0.06 | 7229.9 (17773.8) | 6949.3 (17712.9) | 0.02 |
| **Variables not included in the matching** |  |  |  |  |  |  |
| **Other baseline comorbidities** |  |  |  |  |  |  |
| Cancer | 446 (4.0) | 15953 (5.8) | 0.09 | 442 (4.0) | 470 (4.2) | 0.01 |
| Afib | 172 (1.5) | 11163 (4.1) | 0.15 | 171 (1.5) | 186 (1.7) | 0.01 |
| Glaucoma | 444 (4.0) | 16377 (6.0) | 0.09 | 438 (4.0) | 440 (4.0) | 0.001 |
| MASLD | 655 (5.8) | 14058 (5.1) | 0.03 | 625 (5.6) | 655 (5.9) | 0.01 |
| MASH | 55 (0.5) | 1385 (0.5) | 0.002 | 51 (0.5) | 65 (0.6) | 0.02 |
| Liver fibrosis | 25 (0.2) | 419 (0.2) | 0.02 | 25 (0.2) | 16 (0.1) | 0.02 |
| GERD | 1566 (14.0) | 48361 (17.6) | 0.10 | 1544 (13.9) | 1917 (17.3) | 0.09 |
| Osteoarthritis | 1427 (12.7) | 48113 (17.5) | 0.14 | 1409 (12.7) | 1649 (14.9) | 0.06 |
| Sleep Disorder | 2751 (24.5) | 56551 (20.6) | 0.09 | 2698 (24.4) | 2465 (22.3) | 0.05 |
| PAD | 167 (1.5) | 14450 (5.3) | 0.21 | 163 (1.5) | 267 (2.4) | 0.07 |
| Diabetes Neuropathy | 387 (3.4) | 10895 (4.0) | 0.03 | 375 (3.4) | 498 (4.5) | 0.06 |
| Schizophrenia | 18 (0.2) | 2307 (0.8) | 0.10 | 18 (0.2) | 73 (0.7) | 0.08 |
| Bipolar disorder | 127 (1.1) | 5544 (2.0) | 0.07 | 124 (1.1) | 243 (2.2) | 0.08 |
| Common mental health | 2490 (22.2) | 62832 (22.9) | 0.02 | 2463 (22.2) | 2726 (24.6) | 0.06 |
| Other mental health conditions | 814 (7.3) | 17270 (6.3) | 0.04 | 807 (7.3) | 795 (7.2) | 0.004 |
| **Other baseline medications** |  |  |  |  |  |  |
| Anticoagulant | 227 (2.0) | 13181 (4.8) | 0.15 | 226 (2.0) | 324 (2.9) | 0.06 |
| Antiplatelet | 291 (2.6) | 13536 (4.9) | 0.12 | 284 (2.6) | 448 (4.0) | 0.08 |
| Statin | 3577 (31.9) | 91963 (33.5) | 0.04 | 3487 (31.5) | 3674 (33.2) | 0.04 |
| PCSK9i | 531 (4.7) | 13553 (4.9) | 0.01 | 35 (0.3) | 34 (0.3) | 0.002 |
| Other lipid lowering medications | 37 (0.3) | 1152 (0.4) | 0.02 | 511 (4.6) | 613 (5.5) | 0.04 |
| Sacubitril | 15 (0.1) | 1241 (0.5) | 0.06 | 15 (0.1) | 21 (0.2) | 0.01 |
| Neuropathy medications | 1548 (13.8) | 51526 (18.8) | 0.14 | 1518 (13.7) | 2103 (19.0) | 0.14 |
| Antidepressant | 2944 (26.2) | 76229 (27.8) | 0.04 | 2902 (26.2) | 3337 (30.1) | 0.09 |
| Antipsychotic | 197 (1.8) | 10793 (3.9) | 0.13 | 193 (1.7) | 466 (4.2) | 0.15 |

**Note:** Standardized mean differences (SMDs) were computed as the absolute difference in means (or proportions for categorical variables) between groups divided by the pooled standard deviation. SMDs were calculated using both the TableOne and MatchIt packages to cross-validate covariate balance estimates. To evaluate covariate balance between matched groups, $SMD \leq 0.1$ was used as the criterion for acceptable balance. Unlike $p$ -values, which are influenced by sample size and may indicate statistically significant differences that are not clinically meaningful in large samples, SMD provides a sample size-independent measure of balance that quantifies the magnitude of difference between groups. Therefore, SMDs were used as the primary metric for assessing post-matching balance, while $p$ -values were reported descriptively.
**Abbreviations:** UC, usual care; VINT, Individualized Nutrition Therapy (intervention) program; T2D, type 2 diabetes; MASLD, metabolic dysfunction-associated steatotic liver disease; MASH, metabolic dysfunction-associated steatohepatitis; GERD, gastro-oesophageal reflux disease; Afib, atrial fibrillation; PAD, peripheral artery disease; CKD, chronic kidney disease; SGLT2i, sodium-glucose cotransporter-2 inhibitor; DPP4i, dipeptidyl peptidase-4 inhibitor; RAASi, renin-angiotensin-aldosterone system inhibitor (e.g., ACEi/ARB/ARNI); MRA, mineralocorticoid receptor antagonist (steroidal); non-MRA, non-steroidal mineralocorticoid receptor antagonist (e.g., finerenone); PCSK9i, proprotein convertase subtilisin/kexin type 9 inhibitor.

### Primary Outcomes

During follow-up, incidence rates of new-onset CKD, CKD stage ≥3, and CKD stage ≥4 were lower among VINT participants compared with UC. Incidence rates were 10.1 versus 15.6 per 1,000 person-years for new-onset CKD, 5.7 versus 9.7 per 1,000 person-years for CKD stage ≥3, and 0.5 versus 1.3 per 1,000 person-years for CKD stage ≥4 (Table 3; Figure 2A).

**Figure 2.**
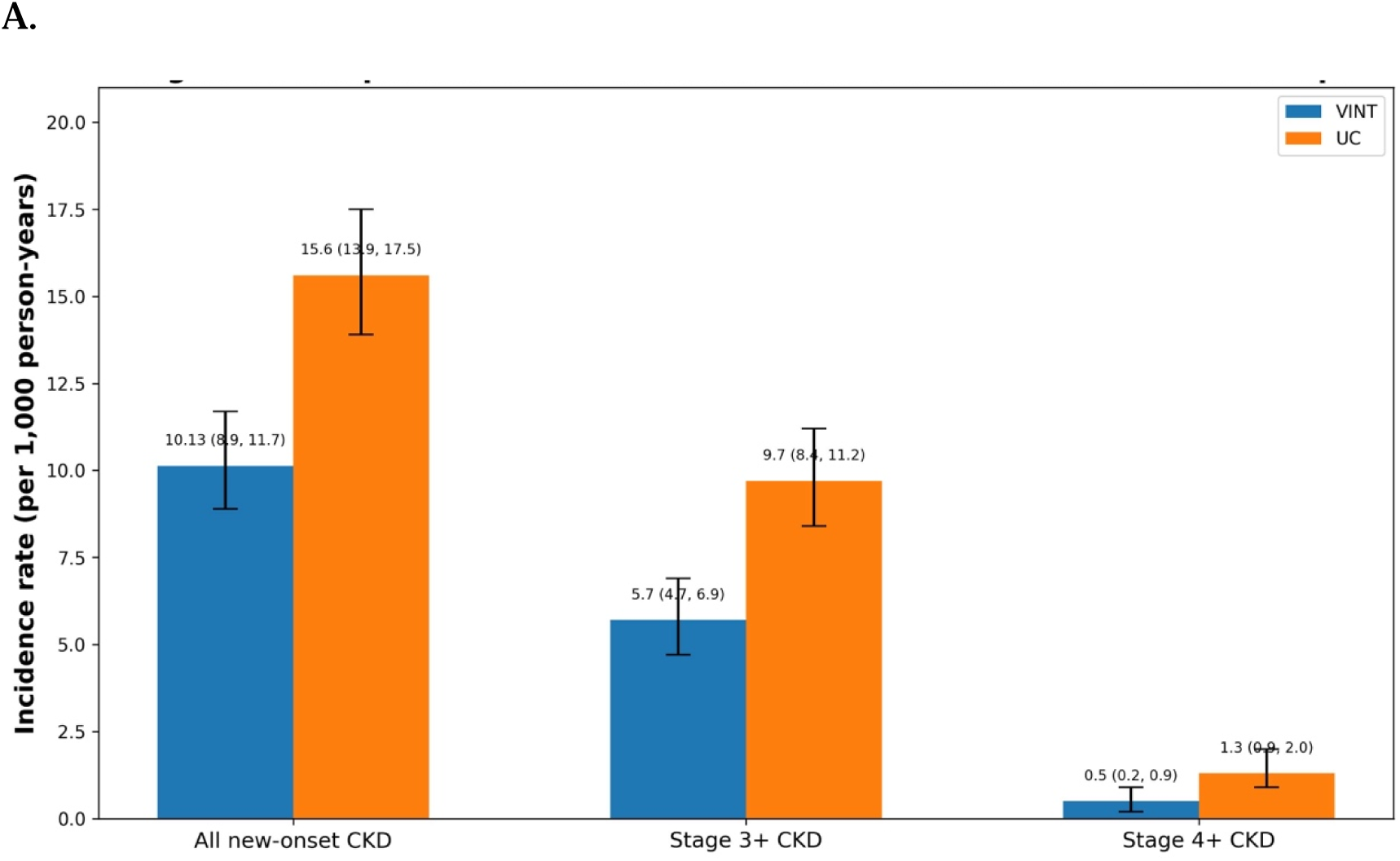
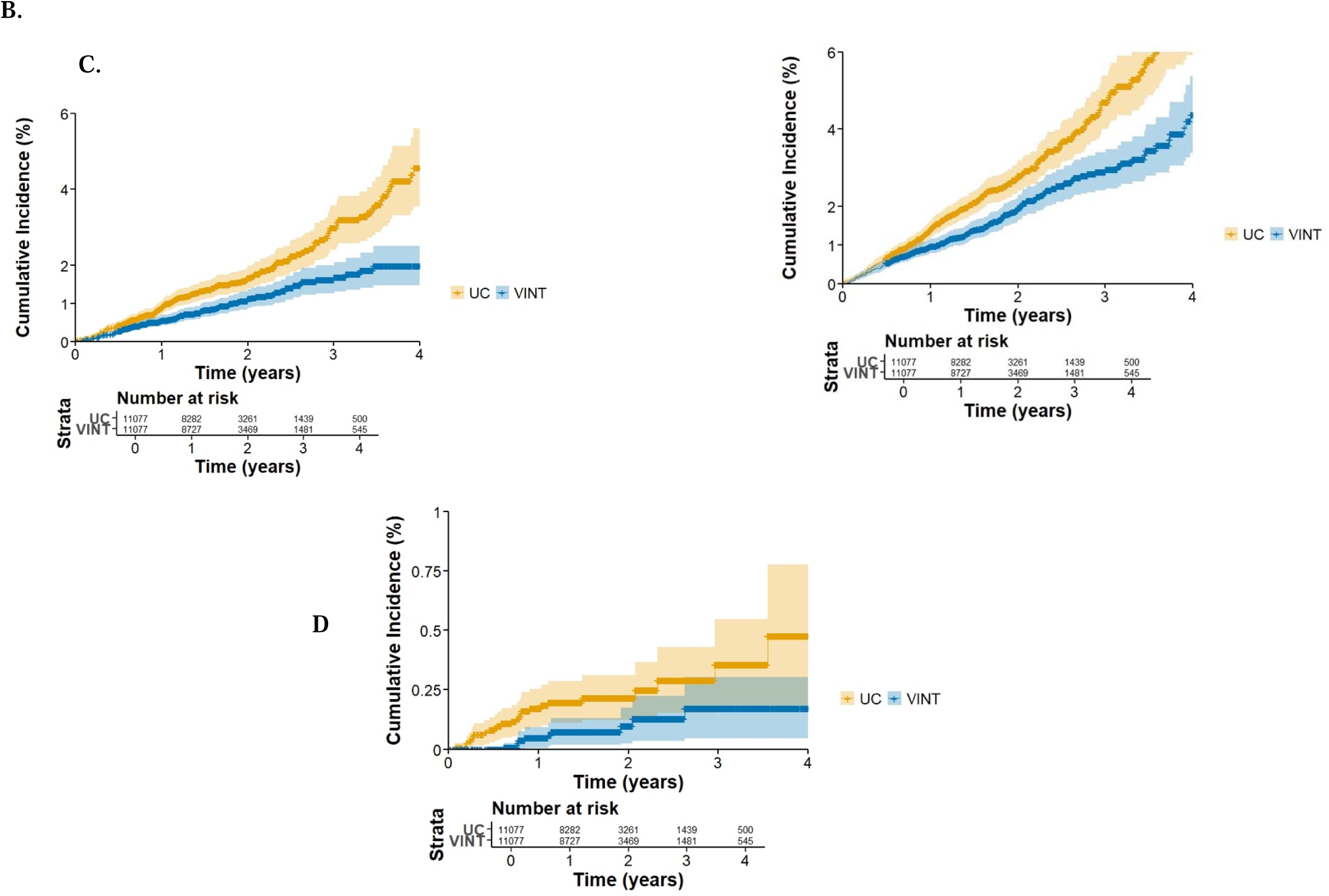
(A) Comparison of CKD Incidence Rates Between VINT and UC Groups Cumulative incidence curves for (B) new-onset CKD (including albuminuria), (C) new-onset stage ≥3 CKD, and (D) new-onset stage ≥4 CKD during follow-up, comparing VINT participants with matched controls (UC). Curves are displayed up to 4 years to aid interpretability, given the decreasing sample size late in follow-up, though all analyses were conducted using the full 5-year observation window with standard right-censoring. At-risk tables are shown below each panel, and steps in the curves reflect incident events. Estimates toward the right tail should be interpreted with caution due to smaller denominators

**Table 3.**
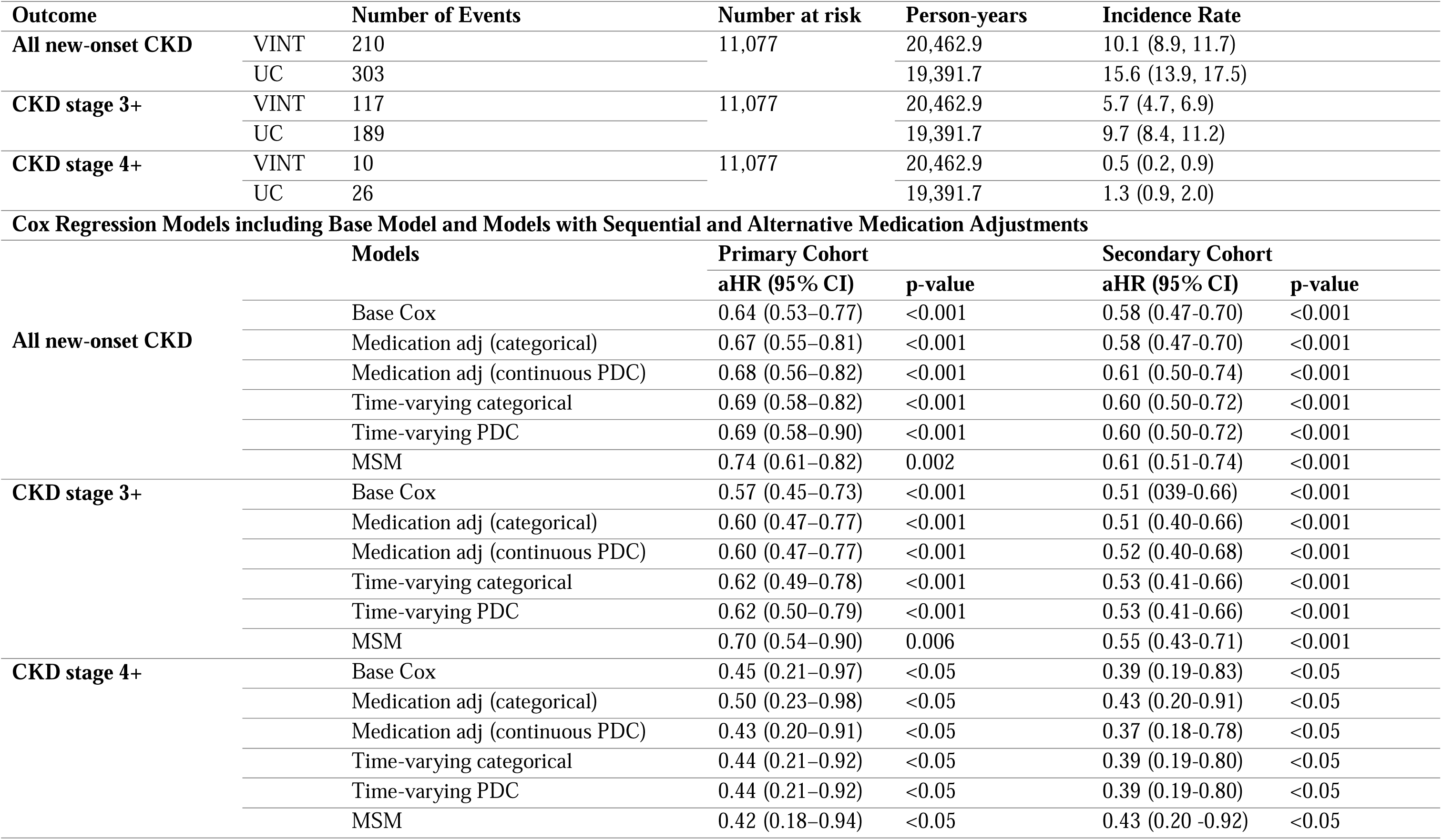
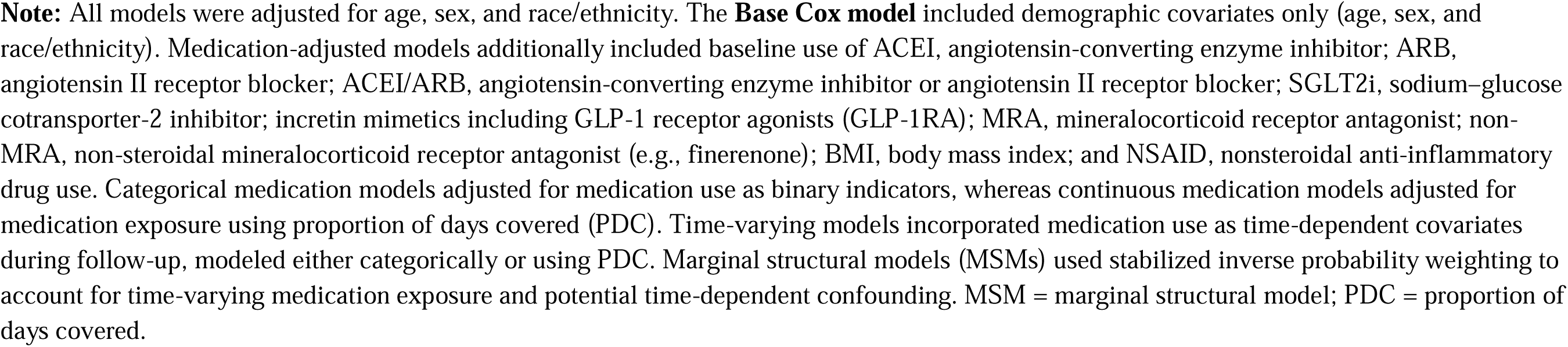
Association of VINT Participation with CKD Outcomes in primary and secondary cohorts.

In Cox proportional hazards models adjusted for age, sex, and race/ethnicity, VINT participation was associated with significantly lower risk of all CKD outcomes compared with UC (Figure 2B-D; Table 3). These associations remained consistent after additional adjustment for medication use, including GLP-1 receptor agonists and SGLT2 inhibitors, regardless of whether medication exposure was modeled as categorical use, proportion of days covered, or time-varying covariates (Table 3; Supplementary Figure S2; Supplementary Table S7). Similarly, in marginal structural models accounting for time-varying confounding and potential mediation by medication use, VINT participation remained significantly associated with lower risk of incident CKD outcomes (Table 3; Supplementary Figure S2). Results were also consistent in Fine–Gray subdistribution hazard models accounting for mortality as a competing event, in which VINT participation remained associated with a significantly lower risk of incident CKD (sHR 0.67, 95% CI 0.55–0.81; p < 0.001). In addition, VINT participation was associated with significantly lower mortality risk compared with matched controls (aHR 0.40, 95% CI 0.21–0.76; p <0.01). The consistency of findings across Cox, marginal structural, and competing-risk models suggests effect estimates were largely unchanged, supporting the robustness of the observed associations.

In a secondary cohort restricted to participants with ≥6 months of VINT participation, associations were similar to and generally stronger than those observed in the primary cohort across all CKD outcomes (Table 3; Supplementary Figure S2; Supplementary Table S7 and S8). Findings remained consistent after adjustment for medication use.

Results from IPTW-weighted Cox models were directionally consistent with the primary analyses, further supporting the robustness of the findings across alternative analytic approaches (Supplementary Tables S9).

### Secondary and exploratory outcomes

Detailed results are provided in the Supplementary Results. In brief, rates of kidney stones, gout, and diabetic ketoacidosis were similar between VINT and usual care, with no statistically significant differences observed. Metabolic acidosis occurred less frequently among VINT participants.

Exploratory analyses suggested that higher mean beta-hydroxybutyrate levels were associated with a lower risk of incident CKD (Supplementary Figure S3), whereas weight loss and HbA1c change were not significantly associated with CKD risk. Among participants with baseline CKD and follow-up claims data, CKD regression or stability occurred more frequently in VINT participants than UC (83.2% vs 73.3%; p = 0.03). In adjusted logistic regression models, VINT participants had a significantly less likelihood of CKD progression compared with controls (OR = 0.51, 95% CI: 0.25–0.99, p = 0.04).

Additional details, including medication use patterns (Supplementary Table S10), are reported in the Supplementary Results.

## Discussion

Our findings demonstrate that participation in the VINT program was significantly associated with a lower risk of incident claims-based CKD compared with a matched control cohort. In this large, longitudinal, claims-based analysis, VINT was associated with a reduced risk of new-onset CKD (∼ 30% reduction), including diagnoses at advanced stages such as stage 3 and beyond (∼ 43% reduction), with a trend toward reduced incidence of stage 4 and beyond (∼ 50% reduction) during follow-up. These results provide novel real-world evidence that a digitally delivered lifestyle intervention emphasizing carbohydrate reduction can meaningfully alter the course of CKD, a condition typically considered progressive and irreversible (27).

Importantly, real-world data analyses such as this extend the relevance of clinical research by capturing outcomes in routine care settings, complementing traditional trials, and informing policy and practice decisions (28). The observation that even advanced-stage CKD diagnoses were prevented, together with findings from exploratory analyses showing a significantly greater likelihood of stage maintenance and regression among participants with baseline CKD in the treated group compared with matched controls, reinforces the potential to improve kidney function in at-risk patients through nutritional and lifestyle interventions.

Telemedicine and digital health interventions, including mobile apps, remote monitoring, and web-based counseling, have been shown to improve diet quality, blood pressure, renal function, and quality of life in adults with CKD (29,30). Beyond clinical outcomes, telehealth improves access, reduces cost and travel burden, and enhances multidisciplinary collaboration, key advantages in CKD care where specialist access is limited. Broader reviews of telenephrology have similarly demonstrated high patient satisfaction, adherence, and scalability (31–33). Integrating these evidence-based digital strategies into a continuous remote care model, the present intervention has previously been shown to reverse and manage T2D and obesity, major CKD risk factors. In this claims-based study, we extend these findings to CKD prevention and progression, suggesting that this approach could be scaled for broader renal care delivery.

The observed reduction in CKD incidence may reflect the unique metabolic effects of nutritional ketosis. Carbohydrate restriction improves insulin sensitivity, inflammation, visceral adiposity, and glycaemic and blood pressure control (19–24, 34), all central to CKD pathogenesis (35). Prior analyses of the same intervention in type 2 diabetes demonstrated stabilization or reversal of eGFR decline (21) and reductions in UACR (24), independent of pharmacologic therapy. In this larger real-world cohort, the protective effect of VINT persisted across multiple modeling approaches, including models that adjusted for medication use as confounders and models that treated medications as mediators, suggesting benefits beyond drug management. Other clinical and real-world evidence supports these findings, with carbohydrate restriction improving eGFR, UACR, and cystatin C (36–40) and lowering risks of kidney failure and mortality (41, 42), whereas high energy-dense carbohydrate intake is associated with a threefold higher CKD risk (43). Unlike traditional low-protein diets, which did not slow CKD progression in the MDRD trial (13, 44), nutritional ketosis targets multiple metabolic pathways simultaneously, offering a more comprehensive disease-modifying strategy.

Consistent with prior studies, participants with mean βHB ≥0.5 mmol/L during the first six months had a significantly lower risk of developing new-onset CKD. In the primary cohort, which included all participants regardless of program duration, effect estimates were modestly attenuated but remained significant, likely reflecting greater heterogeneity in engagement. In contrast, the secondary cohort restricted to ≥6 months of continuous participation showed slightly stronger associations, particularly for more advanced CKD stages, underscoring the importance of sustained exposure. As the ≥6-month definition reflects adherence and metabolic response, these findings are considered exploratory and may be subject to selection or survivor bias. Nevertheless, consistent directionality across cohorts supports the robustness of the association. Sustained carbohydrate restriction, reflected by higher BHB levels, may contribute to these observed benefits. These findings are consistent with prior work demonstrating a dose–response relationship between ketosis and improvements in eGFR slope (21), as well as with Liu et al. (2025), which linked higher βHB levels to improved renal survival and reduced ESRD risk (45). Potential mechanisms include modulation of inflammation, oxidative stress, fibrosis, and renal metabolism (22). Although long-term ketosis trajectories could not be fully assessed, the observed consistency supports the potential role of nutritional ketosis as a kidney-protective strategy, paralleling effects observed with SGLT2 inhibitors (46,47) and supported by evidence from ADPKD studies showing improved eGFR and reduced kidney volume with greater ketosis (48,49).

Concerns regarding kidney-related safety of ketogenic interventions have historically focused on kidney stones (22,50,51) and metabolic acidosis (52,53). In this analysis, kidney stone risk in VINT participants was comparable to matched controls across incidence, prevalence, and recurrence, suggesting no increased risk despite theoretical concerns related to altered urinary calcium, uric acid, and citrate excretion. These findings are consistent with evidence that, under medical supervision with dietary guidance and adequate hydration, nephrolithiasis risk is not elevated (21). Safety outcomes extended beyond nephrolithiasis. Although ketogenic diets are often perceived as high in protein, this intervention employed moderate protein intake, and VINT participants had lower rates of acidosis, potentially reflecting preserved kidney function, particularly in advanced CKD (21,53). Given overlap in ICD-10 coding for acidosis and diabetic ketoacidosis (DKA), DKA was evaluated separately, with no increased risk observed in the VINT group, likely reflecting improved glycemic control and renal function. These findings reinforce that physiologic nutritional ketosis remains distinct from pathologic ketoacidosis (54). Finally, despite concerns regarding gout and renal urate handling, including transient increases during initiation (55), no increase in incident or prevalent gout was observed. Mechanistic evidence further suggests that β-hydroxybutyrate may mitigate gout flares via inhibition of the NLRP3 inflammasome (56).

### Strengths and limitations of study

This study has several strengths, including use of a large external claims’ dataset with up to five years of follow-up, robust propensity score–based matching, and confirmation of findings across complementary analytic approaches, including inverse probability of treatment weighting. Although modest differences in kidney-relevant medication use were observed between cohorts, these therapies were addressed using categorical, continuous, and time-varying models that treated medications as confounders and, in separate analyses, as potential mediators. Associations between the VINT and reduced CKD risk were consistent across analytic approaches, supporting the robustness of the findings.

Several limitations should also be considered. Reliance on closed claims data without linkage to electronic health records or laboratory measures limited follow-up to a subset of the eligible population and precluded direct assessment of kidney function or disease progression using quantitative biomarkers. Outcomes were defined using ICD-10 diagnostic codes, which may introduce misclassification and limit assessment of disease timing, progression, or subclinical changes. Some cases classified as advanced CKD may have reflected pre-existing disease, and analyses focused on incident CKD onset rather than progression of established disease.

Analyses in the secondary cohort, restricted to participants with at least six months of program participation, demonstrated larger effect sizes, suggesting that greater engagement and sustained adherence may enhance the observed benefits of VINT. This extended exposure window also enable exploratory analyses of beta-hydroxybutyrate and weight change within VINT cohort. These analyses were intended to characterize patterns of adherence and metabolic response over time and to inform the design of future studies, rather than to provide causal estimates at enrollment.

Finally, residual confounding related to secular treatment trends, health-seeking behaviors, physical activity, weight loss motivation, digital access, and unmeasured lifestyle or socioeconomic factors cannot be excluded. Dietary intake in the matched control cohort was not captured, and it is therefore unknown whether some individuals in the control group were following similar dietary patterns (e.g., low-carbohydrate or ketogenic diets), which could bias effect estimates toward the null. In addition, VINT represents a multifaceted care model that includes an initial clinical assessment, medication review and adjustment at enrollment, and ongoing medication management in addition to nutritional guidance. Although we incorporated medication-adjusted and time-varying models to account for differences in treatment during follow-up, the independent contribution of baseline clinical assessment and early medication optimization cannot be fully disentangled from the effects of nutritional ketosis. As such, the observed associations likely reflect a combination of metabolic and care-delivery effects. Additionally, the follow-up period may be insufficient to capture longer-term kidney outcomes. No formal sample size calculation was performed, as analyses were exploratory and hypothesis generating. These findings support the need for future randomized and prospective studies integrating claims, electronic health record, and laboratory data to more definitively assess kidney-related effects of nutritional ketosis.

### Conclusions

While RCTs remain the gold standard, our findings provide complementary real-world evidence that extends generalizability to broader, more heterogeneous patient populations and demonstrates effectiveness in routine care settings, offering timely insights to inform clinical guidelines, payer decisions, and health policy (28). These results suggest that the program may offer benefits beyond type 2 diabetes reversal and weight loss, functioning as a comprehensive metabolic intervention with potential relevance to kidney disease risk and progression, an area that merits further investigation. This treatment approach, already established as an evidence-based model for achieving diabetes reversal and sustained weight loss, integrates continuous remote monitoring, medication management, and individualized dietary support focused on carbohydrate restriction. Delivered digitally, this model represents a scalable, nonpharmacologic strategy to help address the growing burden of CKD among metabolically at-risk populations and warrants careful evaluation in patients with more advanced CKD through appropriately designed RCTs.

## Data Availability

The datasets analyzed in this study are not publicly available because they were obtained through a commercial license from the data vendor. Access to these data is restricted, and they were used solely under the terms of the license granted for this study.

## Author contributions

SJA drafted the manuscript, and contributed to the study concept and design, critically reviewed and revised the manuscript, and approved the final version. SJA performed the statistical analyses. SJA and PVS acquired and curated the data. RJJ, PB, AW, TW, JH, and JSV provided clinical and scientific oversight and interpretation of the findings. All authors reviewed and revised the manuscript. RJJ supervised the study and is the guarantor. The corresponding author attests that all listed authors meet authorship criteria and that no others meeting the criteria have been omitted.

## Compliance with Ethics Guidelines

The study was determined to be exempt from institutional review board (IRB) review, as it relied solely on retrospectively collected, de-identified data and did not involve any additional contact or intervention with human subjects.

## Reporting guidelines

This study followed the *Strengthening the Reporting of Observational Studies in Epidemiology (STROBE)* statement for cohort studies. A completed STROBE checklist was prepared and submitted with the manuscript.

## Dissemination

The findings of this study will be disseminated to the public through social media, blogs, and professional networks to enhance accessibility and public understanding. For research transparency and early feedback, the manuscript has been uploaded as a preprint https://www.medrxiv.org/content/10.1101/2025.10.17.25338238v. Once the study is published, we plan to share a summary of the results through digital platforms and academic forums. The study and its findings will also be used to support future research proposals and grant applications aimed at expanding this work and evaluating long-term kidney outcomes in broader populations.

