## Supplemental Material for "Effectiveness of telehealth low-carbohydrate intervention in preventing chronic kidney disease: a real-world, retrospective, matched cohort study"

**Supplementary Method**

**VINT Program Eligibility Criteria**
Participants in the Virta programs (type 2 diabetes reversal and sustainable weight loss) were adults (≥18 years) with overweight, obesity, type 2 diabetes, or prediabetes who were able to engage with program requirements. Eligibility was guided by clinical criteria intended to support the safe initiation and maintenance of nutritional ketosis. These considerations included conditions that may increase clinical risk, such as advanced kidney or liver disease (e.g., end-stage renal disease, or decompensated cirrhosis), pregnancy, certain metabolic or gastrointestinal disorders affecting nutrient absorption, significant electrolyte abnormalities, and serious or unstable psychiatric conditions. In addition, medication use and other clinical factors that could require adjustment or closer monitoring (e.g., insulin or other glucose-lowering therapies, diuretics, or therapies affecting fluid and electrolyte balance) were taken into account.

Importantly, many conditions were not treated as absolute contraindications but rather evaluated in the context of overall clinical stability. Individuals with well-controlled or stable chronic conditions such as chronic kidney disease stage IV-V, compensated liver disease, controlled cardiovascular conditions, or stable psychiatric disorders could be considered appropriate for participation with clinician oversight. In such cases, enrollment was accompanied by individualized care plans, including medication management and ongoing monitoring as needed.

Overall, enrollment reflected real-world clinical practice, where eligibility was determined based on clinician judgment using available medical history and patient readiness, rather than strict application of uniformly verifiable exclusion criteria. As a result, some clinical characteristics that may influence eligibility may not be fully observable in administrative claims data.

**VINT Program’s Nutritional Intervention**

Participants received a nutritional intervention centered on individualized carbohydrate restriction to induce and sustain nutritional ketosis (target β-hydroxybutyrate 0.5–3.0 mmol/L). Protein intake was maintained at moderate levels (1.2–1.5 g/kg reference body weight/day), while fat served as the primary energy source, adjusted according to satiety and energy needs. The dietary approach emphasized whole, minimally processed foods such as non-starchy vegetables, healthy fats (e.g., olive oil, nuts, avocado), and moderate protein sources (e.g., meat, fish, poultry, eggs, dairy), with limited low-glycemic fruit. Processed foods, added sugars, refined grains, and starchy vegetables were discouraged.

Participants were educated on hydration, sodium repletion, and strategies to manage common adaptation effects (e.g., “keto flu”). The intervention was delivered through a digital care platform providing continuous remote support from a multidisciplinary care team, including health coaches and licensed medical providers (e.g., physicians or nurse practitioners). Coaching was delivered using evidence-based behavior change strategies, including goal setting, self-monitoring, motivational interviewing–informed communication, and iterative feedback based on participant progress.

Participants engaged with the platform frequently, particularly during the initial phase of the program. During early participation (e.g., the first 1–3 months), interactions typically occurred daily or near-daily through asynchronous messaging, biomarker reporting, and review of educational content. Over time, the frequency of interaction was individualized based on participant needs, engagement, and clinical stability, with continued access to on-demand support. Synchronous encounters (e.g., video or telephone visits) were conducted as clinically indicated for medical evaluation, medication adjustment, or additional support.

Biomarkers including blood glucose, β-hydroxybutyrate, and body weight were monitored and transmitted through the platform and reviewed by the care team. Monitoring was most intensive during program initiation and was subsequently tailored based on individual progress and tolerance of the intervention. Medication adjustments were performed by clinicians in response to changes in glycemic control, metabolic status, and safety considerations.

Participation in the program was voluntary, and engagement varied across individuals. Participants could reduce their level of interaction or discontinue participation at any time based on personal preference, clinical considerations, or other factors. The program was delivered as a structured, subscription-based telemedicine service; however, specific cost structures varied depending on payer arrangements and are not detailed here. Nutritional recommendations and care plans were continuously adapted to support sustained nutritional ketosis, weight loss, glycemic control, and overall cardiometabolic health.

### **VINT’s Care Team Structure and Participant Support**

Each participant was assigned a dedicated health coach and clinician at enrollment and generally retained the same care team throughout participation in the program. Health coaches provided ongoing education and support related to carbohydrate restriction, nutritional ketosis, self-monitoring, behavior change, goal setting, and problem solving. Coaching interactions were conducted primarily through asynchronous messaging within the digital platform and were informed by evidence-based behavior change techniques, including motivational interviewing, self-monitoring, feedback, and reinforcement strategies.

Clinicians (physicians or nurse practitioners) were responsible for medical management and safety monitoring. Clinical responsibilities included reviewing participant-reported biomarkers and laboratory results, evaluating symptoms and adverse events, adjusting medications, ordering laboratory tests when appropriate, and addressing participant questions related to medical care. Health coaches and clinicians worked collaboratively within the platform to coordinate care and provide timely responses to participant needs.

The frequency and intensity of interactions varied according to participant engagement, clinical complexity, and stage of treatment. Participants had ongoing access to their care team throughout the program and could communicate through the platform as needed. Additional synchronous encounters, including telephone or video visits, were conducted when clinically indicated.

**VINT’s Program Medication Management**

Medication management, led by licensed medical providers, was an integral component of the Virta program to ensure safety during nutritional ketosis and facilitate medication reduction as health improved. At enrollment, participants’ medications were reviewed and individualized deprescription plans were developed. Agents with high hypoglycemia risk, such as insulin and sulfonylureas, were typically reduced or discontinued at initiation, while antihypertensives were closely monitored and adjusted as blood pressure improved. Diuretics were reassessed in the context of carbohydrate restriction–induced natriuresis to prevent volume depletion and electrolyte disturbances.

Other therapies, including GLP-1 receptor agonists, SGLT2 inhibitors, metformin, and lipid-lowering agents, were titrated according to clinical response, tolerability, comorbidities (e.g., CVD or CKD), and laboratory results. Medication management was adaptive: if participants did not maintain carbohydrate restriction or showed worsening metabolic markers, medications were reintroduced or maintained as clinically appropriate. Daily participant-reported biomarkers (blood glucose, ketones, blood pressure, weight) were monitored by the remote care team via the digital platform, allowing real-time communication of adjustments.

This proactive, individualized framework balancing deprescription with reintroduction when needed, minimized risks such as hypoglycemia and hypotension, while enabling systematic evaluation of the independent effects of nutritional ketosis on glycemic control, blood pressure, and other cardiometabolic outcomes

**Baseline demographics, comorbidities and medications**

Baseline comorbidities assessed included type 2 diabetes (T2D), obesity, hypertension, hyperlipidemia, coronary artery disease (CAD), other cardiovascular conditions, stroke, kidney stones, and chronic kidney disease (CKD), including CKD stage and albuminuria. Baseline medication use was identified for renin–angiotensin–aldosterone system inhibitors (RAASi), sodium-glucose cotransporter 2 inhibitors (SGLT2i), incretin mimetics (GLP-1 receptor agonists), dipeptidyl peptidase-4 inhibitors (DPP-4i), insulin, other type 2 diabetes agents, beta-blockers, calcium channel blockers, other antihypertensives, mineralocorticoid receptor antagonists (MRAs), non-mineralocorticoid receptor antagonists, and diuretics. These comorbidities and medication variables, along with demographic characteristics (age, sex, and race/ethnicity) and healthcare costs in the year prior to index/enrollment (pharmacy, inpatient, and outpatient costs), were included as covariates in the propensity score matching model. The ICD-10 codes and medication details are provided in Supplementary Tables S1-S3, respectively. Baseline healthcare costs were also assessed, including total cost of care, pharmacy costs, inpatient costs, and outpatient costs. **Missing baseline data were uncommon (<5% for all covariates). Missing categorical variables were retained as an "Unknown" category, and missing continuous variables were imputed using the median of observed values.**

**Statistical Method**

Categorical variables were summarized using counts and percentages, while continuous variables were summarized using means and standard deviations.

### **Propensity Score Matching**

To reduce baseline differences between VINT participants and usual care controls, propensity score matching was performed using 1:1 nearest-neighbor matching without replacement. Propensity scores were estimated using logistic regression, using a Tier 3 matching strategy with treatment assignment (VINT vs. UC) as the dependent variable.

Covariates included in the propensity score model were:

a) Demographic characteristics (age [coarsened categories with a missingness indicator], sex, race/ethnicity, census region, and area deprivation index quartiles), as well as index year to account for temporal trends.
b) Baseline clinical characteristics included type 2 diabetes, obesity, hypertension, hyperlipidemia, coronary artery disease, other cardiovascular conditions, stroke, and kidney stones. Baseline medication use included renin-angiotensin-aldosterone system (RAAS) inhibitors, SGLT2 inhibitors, incretin mimetics, DPP-4 inhibitors, insulin, other type 2 diabetes therapies, beta-blockers, calcium channel blockers, other antihypertensive agents, mineralocorticoid receptor antagonists, non-mineralocorticoid receptor antagonists, and diuretics. Healthcare utilization and cost variables (prescription drug costs, inpatient costs, and outpatient costs during the year prior to enrollment) were also incorporated to account for baseline healthcare intensity, access to care, and underlying disease burden.

To further strengthen balance and reduce residual confounding, exact matching was enforced on key variables, including sex, age category, diabetes status, obesity status, kidney stone history, select baseline medication classes with renal relevance (e.g., RAAS inhibitors, SGLT2 inhibitors, incretin-based therapies), and index year to ensure temporal alignment between cohorts.

Covariate balance was assessed using standardized mean differences, with an absolute value ≤0.1 considered indicative of acceptable balance.

Baseline characteristics and covariate balance before and after matching are presented in Tables 1 and 2. All variables were selected a priori based on clinical relevance and their potential association with both treatment assignment and CKD outcomes. These variables were comprehensively incorporated into the propensity score model to approximate a counterfactual control population with a similar likelihood of enrolling in VINT if the program had been available. This approach was intended to minimize confounding by indication and ensure comparability between the VINT and usual care cohorts across demographic, clinical, pharmacologic, and healthcare utilization domains.

The primary study outcomes were time to: (1) the first occurrence of a new CKD diagnosis, regardless of stage or albuminuria; (2) the first occurrence of CKD stage 3 or higher (including stages 3, 3a, 3b, 4, 5, ESKD, or initiation of renal dialysis if the diagnosis code was missing); and (3) the first occurrence of CKD stage 4 or higher (including stages 4, 5, ESKD, or initiation of renal dialysis if the diagnosis code was missing) during follow-up. Participants were followed from cohort entry (index date) until the first qualifying CKD event, loss of continuous claims data coverage, or end of study, whichever occurred first.

For stage-specific outcomes, definitions were structured to capture progression to more advanced CKD stages. Individuals who developed lower CKD stages during follow-up (e.g., stage 2 or below for the CKD stage 3+ outcome, and stage 3 or below for the CKD stage 4+ outcome) were censored at the time of occurrence and were not classified as outcome events. This approach ensured that only progression to the specified CKD stage was counted while appropriately accounting for intermediate disease states.

Primary analyses were conducted using Cox proportional hazards regression models applied to the propensity score–matched cohorts. Hazard ratios (HRs) with 95% confidence intervals (CIs) were estimated, and proportional hazards assumptions were evaluated. Robust variance estimators were used to account for the matched design. Incidence rates were calculated as the number of events per 1,000 person-years of follow-up. Person-years were based on total observed follow-up time from the index date until the end of claims availability, disenrollment, or study end and were used for descriptive purposes only.

To account for medication, use during follow-up, models were further adjusted using two complementary approaches: (1) an “ever-on” method, in which medications were coded as binary indicators reflecting any use during follow-up, and (2) a continuous approach based on the proportion of days covered (PDC) for each medication. Medications of interest included SGLT2 inhibitors, incretin mimetics, renin–angiotensin–aldosterone system inhibitors, mineralocorticoid receptor antagonists (steroidal and non-steroidal), diuretics, and nonsteroidal anti-inflammatory drugs (NSAIDs). For medication covariates during follow-up, exposure was determined from available pharmacy claims through the earliest of outcome occurrence, end of claims availability, disenrollment, or study end. The absence of a pharmacy claim during the observation period was assumed to indicate no use of the medication rather than missing data.

To more comprehensively account for time-varying exposure and potential time-dependent confounding, follow-up data were additionally transformed into a time-varying (long-format) dataset structured at the level of calendar quarters. For each participant, follow-up time was divided into sequential quarterly intervals beginning at the index date and continuing until outcome occurrence, loss of coverage, or administrative censoring. Within each interval, medication exposure and covariates were updated based on pharmacy claims. Medication use was parameterized using both binary indicators (present vs absent within a given interval) and continuous measures based on the proportion of days covered (PDC), depending on the modeling approach. In time-varying Cox models, medication exposure was represented using lagged indicators, such that exposure in a given quarter reflected use during the preceding quarter, ensuring appropriate temporal ordering between exposure and outcome.

Time-to-event analyses incorporating time-varying covariates were conducted using Cox proportional hazards models specified with counting-process notation (Surv(tstart, tstop, event)). These models were adjusted for baseline age, sex, race/ethnicity, type 2 diabetes, and obesity, and included clustering at the individual level to account for repeated observations.

In addition to modeling medications as time-varying confounders, marginal structural models (MSMs) were implemented to account for time-dependent confounding affected by prior exposure. Stabilized inverse probability weights were estimated within the quarterly dataset to model the probability of observed medication exposure conditional on prior exposure, baseline covariates, treatment group, and time. Stabilized weights were calculated as the ratio of numerator and denominator probabilities and cumulatively applied across follow-up intervals. To improve numerical stability, weights were truncated at prespecified percentiles. Weighted Cox models were then used to estimate associations between VINT participation and CKD outcomes while treating medication use as a time-varying mediator.

To further evaluate the relationship between sustained exposure and outcomes, a secondary cohort analysis was conducted among participants who remained engaged in the VINT program for at least six months. This analysis was designed to characterize associations within a subgroup with more consistent and prolonged exposure, recognizing that meaningful metabolic and clinical changes may accrue over time. In this cohort, outcome assessment began at program enrollment, consistent with the index date used for the usual care cohort. UC controls were again matched 1:1 to VINT participants using the same propensity score matching framework and stringent matching criteria applied in the primary analysis, including exact matching on race and ethnicity and other key baseline clinical characteristics. Cox proportional hazards models were then used to estimate associations between VINT participation and CKD outcomes.

Finally, a sensitivity analysis was conducted using an inverse probability of treatment weighting (IPTW) approach based on propensity scores estimated from a logistic regression treatment model. The treatment model included demographic, clinical, medication, and health care utilization and cost covariates, consistent with those used in the primary propensity score matching. Propensity score distributions were examined to assess overlap and identify extreme values, and individuals with scores near 0 or 1 were trimmed to improve positivity and reduce the influence of outliers. Stabilized IPTW weights were then calculated and applied in weighted Cox proportional hazards regression models to estimate hazard ratios (HRs) and 95% confidence intervals (CIs) for the three primary CKD endpoints. Covariate balance after weighting was evaluated using standardized mean differences, with absolute values ≤0.1 considered indicative of acceptable balance.

Across analyses, effect estimates from the secondary cohort were compared with those from the primary analysis to assess consistency in direction and magnitude and to evaluate whether longer duration of participation was associated with differences in effect size.

The risk of kidney stone, acidosis, diabetic ketoacidosis, and gout diagnoses was evaluated among participants in the VINT program compared with matched controls over up to five years of follow-up. Outcomes were identified using diagnostic codes consistent with nephrolithiasis (kidney stones) and ICD-10 code E87.2 (acidosis). Analyses were conducted in the full cohort and after excluding participants with a baseline history of the respective condition. Event rates were calculated as the number of diagnoses per 1,000 person-years of follow-up. Group comparisons were performed using unadjusted Poisson regression models with log(person-time) as the offset, reporting incidence rate ratios (IRRs) with 95% CIs. This approach was selected to account for varying follow-up time and to appropriately model recurrent safety events (e.g., kidney stones, acidosis, and gout), which may occur multiple times per individual, especially in the context of safety assessment. Robust standard errors were applied to account for overdispersion. For prevalence analyses, we estimated the proportion of participants with at least one kidney stone diagnosis and the proportion experiencing multiple events during follow-up.

Within the VINT cohort from the secondary matched cohort, exploratory analyses evaluated whether categorical predictors of metabolic response were associated with CKD outcomes. Cox proportional hazards models evaluated time to new CKD onset by ketone category, adjusted first for demographics and then for follow-up medication use (both categorical ever/never and continuous PDC). Incidence rates per 1,000 person-years with 95% CIs were calculated for ketone categories. Predictors included mean ketone levels (≥0.5 mmol/L vs. <0.5 mmol/L), percent weight change at 6 months (≥10% vs. <10%), and A1c change at 6 months or 1 year (≥0.5% vs. <0.5%), modeled as binary variables. Given that mean ketone levels were significant in initial models adjusted for demographics, additional models were estimated with further adjustment for follow-up medication use.

In a subset of participants with baseline CKD stage and available longitudinal follow-up claims data from another set matched VINT and UC baseline CKD and albuminuria, progression and regression of CKD were evaluated, defined as a ≥0 stage decreases or no change in stage during follow-up. Comparisons between groups were performed using Chi-square tests. Logistic regression models estimated odds ratios (ORs) and 95% CIs for the likelihood of progression (regression and stability) in Virta participants compared with controls, adjusted for baseline demographics and follow-up medication use.

Among participants with follow-up CKD diagnoses in the primary 1:1 matched cohort, medication use was further characterized across therapeutic classes, including RAAS inhibitors, mineralocorticoid receptor antagonists (MRAs and non-MRAs), finerenone, incretin mimetics, SGLT2i, diuretics, NSAIDs as well as other diabetes medications. For each class, we summarized the number and proportion of participants with at least one prescription and duration of use, reported as mean and median days supplied. Analyses were stratified by treatment group (VINT vs. UC). Chi-square tests were used to compare proportions, and t-tests compared mean days supplied. Results are presented descriptively as proportions, mean and median duration, and p-values for group comparisons.

### Software

All analyses were conducted in R version 4.3.1 within Komodo Health’s Sentinel environment. Core packages included survival, survminer, MatchIt, cobalt, tableone, gtsummary, broom, ggplot2, sandwich, dplyr, tidyverse, lubridate, tidyr, stringr, reshape2, and purrr. Model outputs were tidied with broom and exported as CSV files for documentation. Statistical analyses were two-sided with an α level of 0.05 and 95% confidence intervals (CIs).

**Supplementary Results**

### **Study Participants Flow**

There were 115,942 VINT participants and 2.39 million potential control participants available in the Komodo Sentinel environment for this claims-based analysis (Figure 1; Supplementary Table S4). After applying data availability requirements, including at least one year of pre-enrollment/index data and continuous closed claims coverage, 14,376 VINT participants and 727,030 control participants remained eligible. Participants with severe comorbid conditions, including ESKD/dialysis, active or metastatic cancer, transplant status, hospice or critical care utilization, and advanced organ failure, were subsequently excluded. Additional exclusions included type 1 diabetes, age <18 years, insufficient follow-up (<6 months of claims data), and non-CKD kidney diseases or advanced renal conditions that could independently influence CKD outcomes (Figure 1; Supplementary Table S4).

To ensure that only incident CKD outcomes were evaluated, participants with baseline CKD or albuminuria were excluded prior to matching. After all eligibility criteria and exclusions were applied, 11,225 VINT participants and 274,455 control participants remained eligible for the primary analysis. A Tier 3 (most stringent) 1:1 propensity score matching approach was then used to construct a control cohort with a similar likelihood of enrolling in VINT. The matching algorithm incorporated demographic characteristics, index year, baseline comorbidities, disease severity, medication use, healthcare utilization, and socioeconomic variables, with exact matching on key clinical characteristics. The final matched analytic cohort consisted of 11,077 VINT participants and 11,077 matched usual care controls.

### **Baseline Characteristics**

Following 1:1 propensity score matching, the final analytic sample included 11,077 VINT participants and 11,077 matched usual care (UC) controls (Figure 1; Tables 1 and 2). To evaluate representativeness, the matched VINT cohort was compared with VINT participants who were excluded during cohort selection (Supplementary Table S5). Baseline demographic and clinical characteristics were generally similar between the included and excluded VINT cohorts, with most standardized mean differences (SMDs) <0.1. Larger differences were observed for race/ethnicity and enrollment year, variables that were subsequently addressed through the matching process.

### In the final matched cohort, baseline characteristics were well balanced between VINT and UC participants, with all matching variables demonstrating SMDs <0.1 (Tables 1 and 2). The mean age was 51 years, and approximately 60% of participants were female. By design, all participants were free of baseline CKD and albuminuria. The prevalence of major comorbidities included type 2 diabetes (36%), obesity (55%), hypertension (42%), and hyperlipidemia (44%). Mean follow-up was 1.9 years in the VINT cohort and 1.8 years in the UC cohort. Control index dates were aligned to the VINT enrollment distribution to account for temporal trends and calendar-time effects (Supplementary Figure S1). Similarly, baseline characteristics in the secondary matched cohort were also well balanced between groups, with all SMDs <0.1 (Supplementary Table S6).

**Primary Outcomes**

Primary CKD outcomes are presented in results section of the main document with reference to Table 3 and Supplementary Tables S7–S9.

### **Secondary Safety Outcomes**

Over 5 years of follow-up, the incidence rate of kidney stone diagnoses was 14.7 per 1,000 person-years among VINT participants and 16.3 per 1,000 person-years among UC. After excluding individuals with a history of kidney stones at baseline, incidence rates were 11.3 versus 12.6 per 1,000 person-years, respectively. In unadjusted Poisson regression models, there was no statistically significant difference in kidney stone risk between groups (all participants: IRR = 0.90, 95% CI 0.77–1.06, p = 0.20; excluding baseline cases: IRR = 0.90, 95% CI 0.75–1.07, p = 0.23). Prevalence analyses showed similar findings, with kidney stones diagnosed in 2.0% of VINT participants and 2.2% of UC participants during follow-up. After excluding participants with baseline kidney stones, prevalence was 1.7% and 1.8%, respectively. Multiple kidney stone events were uncommon and occurred at similar frequencies in VINT and UC participants (0.47% vs. 0.48%, respectively; 0.34% vs. 0.33% after excluding baseline cases).

Given the non-specific nature of ICD-10-CM E87.2 (metabolic acidosis), we prespecified acidosis and diabetic ketoacidosis (DKA) as a safety domain and report each outcome separately. The incidence of metabolic acidosis was 2.7 per 1,000 person-years in VINT participants compared with 7.2 per 1,000 person-years in UC. In unadjusted Poisson regression models, VINT participation was associated with a significantly lower incidence of acidosis (IRR = 0.38, 95% CI 0.27–0.51; p < 0.001). DKA incidence was 5.6 per 1,000 person-years in the VINT group and 7.5 per 1,000 person-years in UC. In Cox proportional hazards models, VINT participation was not significantly associated with incident DKA (HR 0.79, 95% CI 0.61–1.02; p = 0.075).

The prevalence of gout was 1.9% among VINT participants and 2.2% among UC participants during follow-up. Incidence rates were 17.6 and 20.9 per 1,000 person-years, respectively. In unadjusted Poisson regression models, VINT participation was not significantly associated with incident gout (IRR = 0.84, 95% CI 0.74–1.07; p = 0.054), indicating no meaningful difference in gout risk between groups.

### **Exploratory Outcomes**

Among matched VINT participants with available biomarker data (Supplementary Table S5), 6-month mean βHB concentrations were distributed as follows: 28.0% had mean βHB levels of 0–0.3 mmol/L, 30.5% had levels of 0.3–0.5 mmol/L, 31.6% had levels of 0.5–1.0 mmol/L, and 9.8% had levels ≥1.0 mmol/L. For 6-month weight change, 34.0% of participants lost <5% of baseline body weight, 27.3% lost 5–<10%, 21.2% lost 10–<15%, 11.4% lost 15–<20%, and 6.1% lost ≥20% of baseline body weight.

Per the analysis plan within the VINT participants from the secondary cohort, missing βHB values were grouped with <0.5 mM; aggregating categories, <0.5 mM (including missing) accounted for 58.0%, and ≥0.5 mM for 42.0%. In Cox proportional hazards models, mean BHB levels ≥0.5 mmol/L over 6 months were associated with a significantly lower risk of CKD in models adjusted for demographics (HR = 0.69, 95% CI: 0.50–0.95, p = 0.02; Figure 5). This association remained consistent after additional adjustment for medication use (HR = 0.75, 95% CI: 0.54–0.98, p = 0.04). Incidence rates of CKD were 10.9 per 1,000 person-years (95% CI: 9.0–13.0) among participants with mean BHB levels <0.5 mmol/L compared with 9.4 per 1,000 person-years (95% CI: 7.4–11.8) among those with mean BHB levels ≥0.5 mmol/L. By contrast, neither 6-month weight change (≥10% vs <10%; HR, 0.79; 95% CI, 0.57–1.11; p=0.18) nor HbA1c change at 6 or 12 months (≥0.5 percentage points vs <0.5; HR, 0.94; 95% CI, 0.63–1.40; p=0.76) was significantly associated with incident CKD.

Among participants with baseline CKD stage and available follow-up claims data (n =271), 104 of 125 VINT participants (83.2%) demonstrated regression or stability of CKD diagnosis compared with 107 of 146 UC (73.3%). This difference was statistically significant (p = 0.03). In adjusted logistic regression models, Virta participants had a significantly less likelihood of CKD progression compared with controls (OR = 0.51, 95% CI: 0.25–0.99, p = 0.04).

**Medication Use and Persistence**

Medication use among participants with a CKD diagnosis during follow-up who had available prescription claims data (n = 210 VINT and n = 303 UC) was generally comparable between groups across most major therapeutic classes (Supplementary Table S10). Use of GLP-1 receptor agonists was significantly higher among VINT participants compared with UC (58.6% vs. 41.7%, p = 0.0002), whereas SGLT2 inhibitor use was similar between groups (41.4% vs. 33.0%, p = 0.064). Use of RAAS inhibitors, diuretics, mineralocorticoid receptor antagonists (MRA/non-MRA), finerenone, and NSAIDs did not differ significantly between groups.

Patterns of medication exposure, assessed by days of use, were broadly similar between groups. However, VINT participants had significantly greater exposure to RAAS inhibitors (mean 1,217 vs. 1,015 days, p = 0.034), whereas exposure to GLP-1 receptor agonists showed a similar trend that did not reach statistical significance (868 vs. 704 days, p = 0.054). No significant differences were observed in duration of use for SGLT2 inhibitors, diuretics, MRA/non-MRA therapies, or NSAIDs. Finerenone use was infrequent in both groups (2.9% vs. 2.3%, p = 0.93), precluding meaningful comparisons of treatment duration.

**STROBE Statement—Checklist**

| Section | Item No | Recommendation | Page No | Addressed in Manuscript |
| --- | --- | --- | --- | --- |
| **Title and abstract** | 1(a) | Indicate the study’s design with a commonly used term in the title or the abstract | 1 | Title specifies “retrospective, propensity score–matched cohort study.” |
|  | 1(b) | Provide in the abstract an informative and balanced summary of what was done and what was found | 2 | Abstract summarizes objectives, design, setting, participants, main outcomes, results, and conclusions. |
| **Introduction** | 2 | Explain the scientific background and rationale for the investigation being reported | 4-5 | Background outlines CKD burden, rationale for individualized nutrition therapy in type 2 diabetes and obesity. |
|  | 3 | State specific objectives, including any prespecified hypotheses | 5 | Objective clearly stated: to evaluate the effectiveness of a telehealth individualized nutrition therapy (VINT) for CKD prevention and progression. |
| **Methods** | 4 | Present key elements of study design early in the paper | 5 | Retrospective matched cohort design using the Komodo Healthcare Map™ claims database. |
|  | 5 | Describe the setting, locations, and relevant dates, including periods of recruitment, exposure, follow-up, and data collection | 7 | U.S.-based claims dataset; enrollment from 2015–2025; up to 5 years of follow-up. |
|  | 6(a) | Give the eligibility criteria, and the sources and methods of selection of participants. Describe methods of follow-up | 7 and supplementary method | Inclusion/exclusion criteria and continuous claims follow-up detailed in Methods. |
|  | 6(b) | For matched studies, give matching criteria and number of exposed and unexposed | 8-10, supplementary method | 1:1 propensity score matching on demographics, index year, comorbidities, and medication covariates; sensitivity analyses using IPTW. |
|  | 7 | Clearly define all outcomes, exposures, predictors, potential confounders, and effect modifiers | 7–8, supplementary method | Outcomes (CKD onset, stages ≥3/≥4, safety, regression/stability) and confounders defined. |
|  | 8* | For each variable of interest, give data sources and methods of assessment | 6-7, supplementary method | ICD-10-CM, CPT/HCPCS codes, and pharmacy claims used; same coding in both groups. |
|  | 9 | Describe efforts to address potential sources of bias | 8-10, supplementary method | Propensity matching, IPTW weighting, and covariate adjustment used to reduce confounding. |
|  | 10 | Explain how study size was arrived at | 10-11 | All eligible participants meeting inclusion criteria included 11,077 VINT vs 11,077 UC matched pairs. |
|  | 11 | Explain how quantitative variables were handled in analyses | 8-10 | Continuous variables summarized as mean (SD); categorical cutoffs for A1c, BHB, weight loss; SMDs used for covariate balance. |
|  | 12(a) | Describe all statistical methods, including those used to control for confounding | 8–10 | Cox proportional hazards and Poisson regression adjusted for demographics, comorbidities, and medications. |
|  | 12(b) | Describe any methods used to examine subgroups and interactions | 8-10 | Subgroup analyses for BHB, weight change, and A1c categories using Cox and GLM models. |
|  | 12(c) | Explain how missing data were addressed | 8, 11 | Primary and secondary outcomes were evaluated using survival models that included all participants with available claims follow-up. Participants contributed person-time until outcome occurrence or censoring, allowing for variable follow-up durations. For BHB, HbA1c, and weight change, missing values were treated as not achieving the target and handled using a worst-observation-carried-forward (WOCF) approach by assigning them to the lowest exposure or least improvement category. Missing covariate values were categorized as unknown. |
|  | 12(d) | Explain how loss to follow-up was addressed | 8 | Participants censored at loss of continuous claims coverage, when event occurred or end of observation. |
|  | 12(e) | Describe any sensitivity analyses | 8–10, supplementary method | IPTW analyses conducted; results consistent across models. Models using medication as time varying covariates and mediators |
| **Results** | 13(a) | Report numbers of individuals at each stage of study | 10 | Figure 1 and Supplementary Table S3 shows participant flow: from 2.39 million controls to final 11,366 matched pairs. |
|  | 13(b) | Give reasons for non-participation at each stage | 10 | Exclusions for missing data, incomplete claims, or safety conditions explained. |
|  | 13(c) | Consider use of a flow diagram | 10 | Figure 1 provides study flow diagram. |
|  | 14(a) | Give characteristics of study participants and confounders | 10–11 | Table 1: demographics, comorbidities, medication use; all covariates balanced (SMD <0.1). |
|  | 14(b) | Indicate number of participants with missing data for each variable | 11, supplementary table S3 | Missingness reported for BHB, A1c, and weight change. |
|  | 14(c) | Summarise follow-up time | 10 | Mean follow-up 1.9 years (VINT) vs 1.8 years (UC). |
|  | 15* | Report numbers of outcome events or summary measures over time | 11–14 | CKD incidence, stage transitions, regression, and safety outcomes with HRs and IRRs. |
|  | 16(a) | Give unadjusted and adjusted estimates and their precision | 11–14 | HRs with 95% CIs and p-values for unadjusted and adjusted models. |
|  | 16(b) | Report category boundaries when continuous variables categorized | 11–13 | BHB ≥0.5 mmol/L, weight loss ≥10%, A1c change ≥0.5%. |
|  | 16(c) | Translate estimates of relative risk into absolute risk if relevant | 11-14 | Incidence rates per 1,000 person-years presented. |
|  | 17 | Report other analyses done | 13–14 | Sensitivity and exploratory analyses on BHB, weight, and A1c included. |
| **Discussion** | 18 | Summarise key results with reference to objectives | 14–15 | 30-55% lower CKD incidence in VINT vs UC summarized. |
|  | 19 | Discuss limitations, potential bias, or imprecision | 19 | ICD misclassification, residual confounding, and follow-up limitations discussed. |
|  | 20 | Give cautious overall interpretation | 18–20 | Balanced interpretation referencing consistency with prior work and biological plausibility. |
|  | 21 | Discuss generalisability (external validity) | 20 | Generalizable to real-world U.S. adults with T2D, obesity and overweight in claims datasets. |
| **Other information** | 22 | Give source of funding and role of funders | 34 | No external funding; retrospective analysis of deidentified claims data. |

*Information provided separately for exposed and unexposed (VINT vs UC) groups.*

**Summary:** All STROBE checklist items were addressed in the manuscript and/or supplementary materials. The study transparently reports design, data source, matching, statistical methods, sensitivity analyses, and interpretation according to STROBE standards.

**Supplementary Figure S1.** Distribution of index dates among VINT participants and matched UC controls. Bars represent the count of participants initiating follow-up on a given index date, plotted separately for VINT (red) and UC (blue). The figure illustrates the temporal overlap in cohort enrollment, showing increasing accrual over time with peaks corresponding to more recent years. The overlap between VINT and UC participants across the study period reduces the potential for confounding by calendar time or seasonal effects.

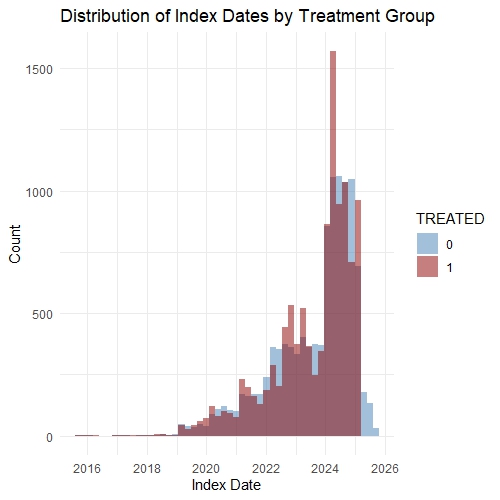

**Supplementary Figure S2. Risk of CKD outcomes by model adjustment.**
Forest plot of hazard ratios (HRs) and 95% confidence intervals for new-onset chronic kidney disease (CKD), CKD stage 3 or higher, and CKD stage 4 or higher among VINT participants compared with matched UC) controls in the primary and secondary cohorts. Estimates are shown across different adjusted Cox proportional hazards models, including adjustment for demographics only, categorical medication use, continuous medication exposure modeled as proportion of days covered (PDC), and time-varying medication exposure, as well as marginal structural models (MSMs) accounting for time-varying confounding. Points indicate HRs and horizontal lines indicate 95% confidence intervals; the dashed vertical line denotes the null value (HR = 1.0).

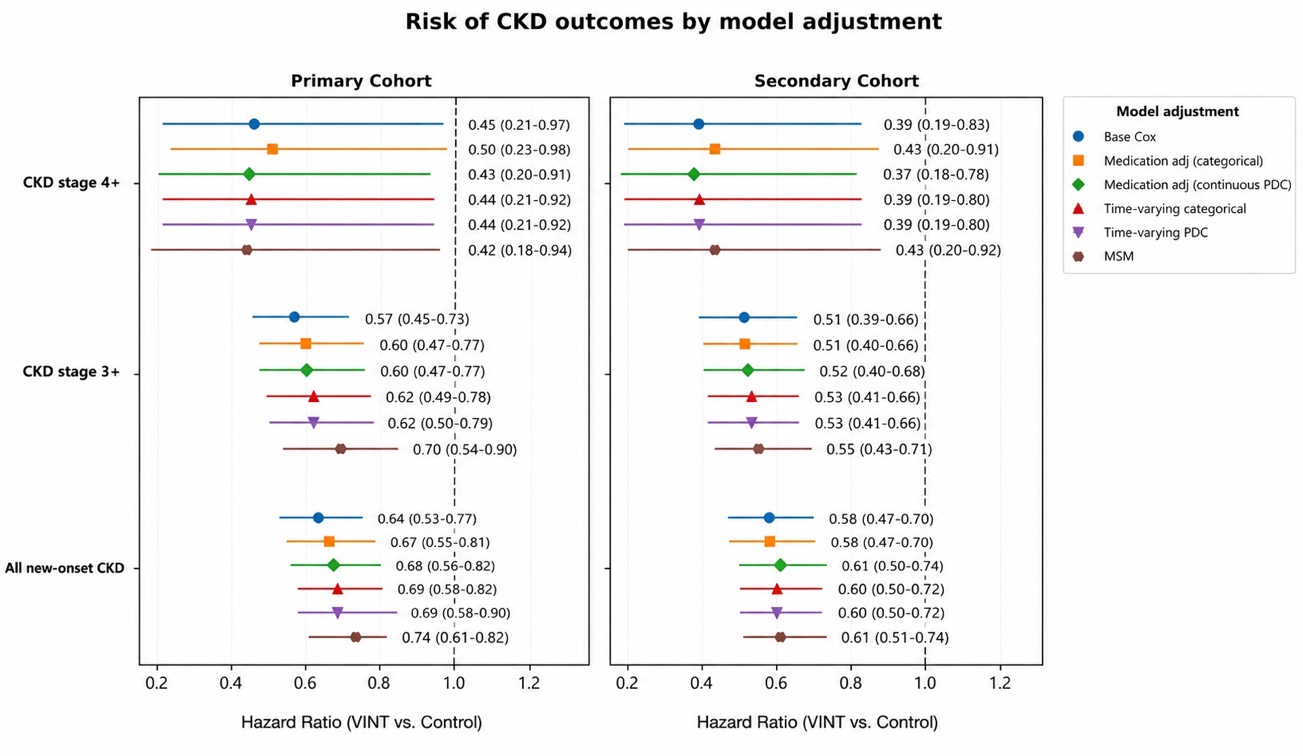

**Supplementary Figure S3.** Cumulative incidence curves for new-onset CKD during follow-up stratified by mean 6-month beta-hydroxybutyrate (BHB) concentration (<0.5 mmol/L vs. ≥0.5 mmol/L). Participants with CKD at baseline were excluded. Curves are displayed through 4 years to improve visualization, although all analyses were conducted using the full 5-year follow-up period with standard right-censoring. Shaded areas represent 95% confidence intervals. Numbers at risk are shown below each panel. Estimates at later time points should be interpreted with caution because of the decreasing number of participants remaining under observation.

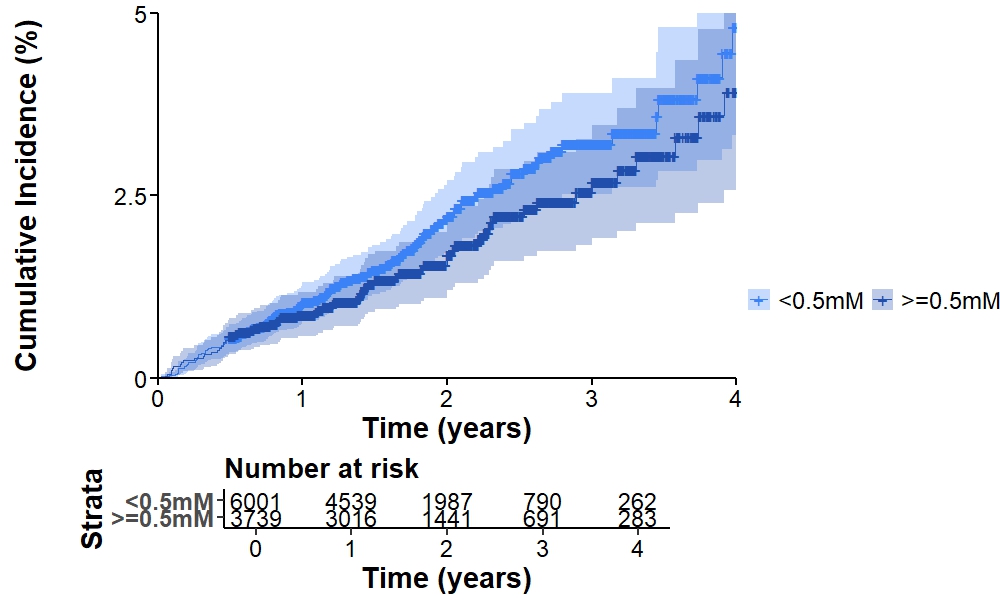

**Supplementary Table S1.** ICD-10 code sets for identifying primary, exploratory and safety outcomes assessed in the study

| **ICD-10** | **Description** |
| --- | --- |
| **Primary Outcome** | |
| 1) **All new CKD diagnosss, including albuminuria** | |
| N18.1 | Stage 1 CKD |
| N18.2 | Stage 2 CKD |
| N18.3 including N18.31, N18.32 | Stage 3 CKD |
| N18.4 | Stage 4 CKD |
| N18.5 | Stage 5 CKD |
| N18.6 | End Stage Kidney Disease |
| Z992 | Renal dialysis |
| Z49 | Renal dialysis |
| R80 | Albuminuria |
| 2) **Stage ≥3 CKD diagnosis** | |
| N18.3 including N18.31, N18.32 Stage 3 CKD | |
| N18.4 Stage 4 CKD | |
| N18.5 Stage 5 CKD | |
| N18.6 End Stage Kidney Disease | |
| Z992 Renal dialysis | |
| Z49 Renal dialysis | |
| **Exploratory Outcome** | |
| **Stage ≥4 CKD diagnosis** | |
| N18.4 Stage 4 CKD | |
| N18.5 Stage 5 CKD | |
| N18.6 End Stage Kidney Disease | |
| Z992 Renal dialysis | |
| Z49 Renal dialysis | |
| **Safety outcomes** | |
| N20 | Any Kidney Stones |
| E87.2 | Any Acidosis |
| M10, M1A | Acute and chronic gout |
| E131, E1310, E1311 | Other specified diabetes mellitus with ketoacidosis, with or without coma |
| E111, E1110, E1111 | Type 2 diabetes mellitus with ketoacidosis, with or without coma |

**Supplementary Table S2.** ICD-10 code sets for identifying baseline comorbidities and other follow-up comorbidities used for matching or matching assessment

| **ICD-10** | **Description** |
| --- | --- |
| **Exclusion Criteria** | |
| Z515 Hospice palliative | |
| Q5001-Q5010, G0182 Hospice HCPS | |
| A40, A41, R652. Sepsis (critical care) | |
| 94002-94003, 94656-94657, E0465-E0467 Respiratory Support | |
| J44, J43, J96. COPD- Respiratory | |
| 33979-33981, 33990-33993. Heart failure Support | |
| Z94 Transplant status | |
| Z51, Z5111. Active cancer treatment | |
| C77-C80 Metastatic cancer | |
| K70, B18, K753, K754. Liver exclusion | |
| **Exclusion Kidney Disease (excluded baseline)** | |
| Q60 - 63 | Congenital malformations of the urinary system |
| N00-N08 | Glomerular diseases |
| N10-N17 | Renal tubule-interstitial diseases, Acute kidney injury |
| M32.14 | Lupus nephritis |
| C64-C65 | Malignant neoplasms of kidney and renal pelvis |
| N13 | Obstructive and reflux uropathy |
| N20-N23 | Urolithiasis |
| N28 | Non-specific renal disease |
| Z992, Z49 | Renal Dialysis |
| Procedure codes 0TY00Z, 5A1D | Kidney transplant and dialysis procedures |
| CPT codes 90935-90947 | Hemodialysis procedures |
| **Baseline Comorbidities** | |
| E11 or E13 | Type 2 diabetes |
| E10 | Type 1 diabetes |
| E66 | Obesity and BMI 30+ |
| E78 | Hyperlipidemia |
| Hypertension | |
| I10 | Essential |
| I11 | Hypertensive heart disease |
| I12 | Hypertensive chronic kidney disease |
| I13 | Hypertensive heart & chronic kidney disease |
| I15.0,2,9 | Secondary hypertension |
| I16.0, 1 | Hypertensive crisis |
| Z720 | Smoking |
| C* | Any cancer |
| O* or Z3* | Pregnancy |
| Coronary Artery Disease (CAD) | |
| Ischemic Heart Disease (I20, I23, I24, I25) | |
| I24 | Acute ischemic heart disease |
| I25 | Chronic ischemic heart disease |
| I20 | Preinfarction syndrome |
| I25 | Aneurysm of coronary vessels |
| I20.1/8/9 | Angina pectoris |
| I25.81 | Arteriosclerosis of coronary artery bypass graft |
| I25.10 | Atherosclerosis of coronary artery without angina pectoris |
| I24.0 | Coronary thrombosis not resulting in myocardial infarction |
| I25.6 | Silent myocardial ischemia |
| I25.111 | Coronary artery spasm |
| I25.82 | Chronic total occlusion of coronary artery |
| I20.0 | Unstable angina co-occurrent and due to coronary arteriosclerosis |
| I25.119 | Angina co-occurrent and due to coronary arteriosclerosis |
| I25.42 | Dissection of coronary artery |
| I25.811 | Coronary arteriosclerosis in artery of transplanted heart |
| I25.812 | Arteriosclerosis of coronary artery bypass graft of transplanted heart |
| I25.1 | Coronary atherosclerosis |
| I25.719 | Arteriosclerosis of autologous vein coronary artery bypass graft |
| I25.739 | Arteriosclerosis of nonautologous coronary artery bypass graft |
| I25.729 | Arteriosclerosis of autologous arterial coronary artery bypass graft |
| I23.7 | Post infarct angina |
| I23.1 | Atrial septal defect due to and following acute myocardial infraction |
| I25.5 | Generalized ischemic myocardial dysfunction |
| I23.0,3,4,5,6 | Rupture of chordae tendinae due to and following acute myocardial infarction |
|  | Rupture of papillary muscle as current complication following acute myocardial infraction |
|  | Hemopericardium due to and following acute myocardial infraction |
|  | Rupture of cardiac wall without hemopericardium as current complication following acute myocardial infarction |
|  | Thrombosis of atrium, auricular appendage, and ventricle due to and following acute myocardial infraction |
| Heart attack (I21) | |
| I21.X | Acute non-ST segment elevation myocardial infarction |
|  | Acute ST segment elevation myocardial infraction |
|  | Myocardial infarction |
|  | Myocardial infarction due to demand ischemia |
|  | Acute ST segment elevation myocardial infarction involving left anterior descending coronary artery |
|  | Acute ST segment elevation myocardial infarction due to left coronary artery occlusion |
|  | Acute ST segment elevation myocardial infarction due to right coronary artery occlusion |
| I22.X | Subsequent STEMI of anterior wall |
|  | Subsequent STEMI of interior wall |
|  | Subsequent STEMI of other sites |
|  | Subsequent MI, unspecified |
| Heart Failure (I50) | |
| I50.X | Congestive heart failure |
|  | Left heart failure |
|  | Systolic heart failure |
|  | Diastolic heart failure |
|  | Hypertensive heart failure |
|  | Chronic right-sided heart failure |
|  | Biventricular congestive heart failure |
|  | Acute on chronic right-sided congestive heart failure |
|  | Chronic systolic heart failure |
|  | Chronic diastolic heart failure |
|  | Acute on chronic systolic heart failure |
|  | Acute on chronic diastolic heart failure |
|  | Chronic combined systolic and diastolic heart failure |
|  | Acute on chronic combined systolic and diastolic heart failure |
|  | Acute right-sided heart failure |
|  | Acute systolic heart failure |
|  | Acute diastolic heart failure |
|  | Acute combined systolic and diastolic heart failure |
|  | Heart Failure |
| Peripheral Vascular Diseases (PAD) | |
| I70, 73 | Peripheral vascular disease |
| Composite Cerebrovascular | |
| Stroke (I60-I64) | Subarachnoid hemorrhage |
|  | Intracerebral hemorrhage |
|  | Other nontraumatic intracranial hemorrhage |
|  | Cerebral infarction |
|  | Stroke, not specified as hemorrhage or infraction |
| Transient Ischemic Attack (G45.x) | Transient cerebral ischemic attacks and related syndromes |
| MASLD/MASH | |
| K76, K760, K7685 | MASLD |
| K758-K7581 | MASH |
| I48.X | Afib |
| G47 | Sleep disorder |
| M15-M19 | Osteoarthritis |
| K21 | GERD |
| H40 | Glaucoma |

**Supplementary Table S3.** List of drug names used to identify prescription drug use at baseline and during follow-up

| Drug category | Generic names |
| --- | --- |
| SGLT2i | Canagliflozin  Dapagliflozin  Empagliflozin  Ertugliflozin  Sotagliflozin |
| Sulfonylureas | Glipizide  Glyburide  Glimepiride  Chorpropamide  Tolbutamide  Tolazamide  Acetohexamide |
| DPP4 | Sitagliptin  Saxagliptin  Linagliptin  Alogliptin  Vildagliptin  Teneligliptin |
| Thiazoledinedione | Pioglitazone  Rosiglitazone |
| Insulin | Insulin  Humulin  Humalog  Novolog |
| GLP-1 | Albiglutide  Exenatide  Liraglutide  Lixisenatide  Dulaglutide  Semaglutide  Tirzepatide |
| Other T2D medications | Metformin  Acabose  Miglitol  Repaglinide  Nateglinide  Pramlintide |
| Anticoagulant | Warfarin  Dabigatran  Rivaroxaban  Apixaban  Edoxaban  Betrixaban  Enoxaparin  Heparin  Fondaparinux |
| Antiplatelets | Anagrelide  Aspirin  Abciximab  Cilostazol  Clopidogrel  Dipyridamole  Eptifibatide  Prasugrel  Icagrelor  Ticlopidine  Vorapaxar  Cangrelor  Tirofiban |
| Statins | Atorvastatin  Simvastatin  Rosuvastatin  Pravastatin  Lovastatin  Fluvastatin  Pitavastatin |
| Other lipid lowering medications | Ezetimibe  Cholestyramine  Colestipol  Colesevelam  Fenofibrate  Gemfibrozil  Niacin  Icosapent ethyl  Omega-3 acid esters |
| PCSK9i | Alirocumab  Evolocumab  Inclisiran |
| MRA | Eplerenone  Spironolactone |
| Non MRA | Finerenone |
| Diuretic | Bendroflumethiazide  Chlorothiazide  Chlorthalidone  Hydrochlorothiazide  Indapamide  Myethoclothiazide  Metolazone  Bumetanide  Ethacrynate sodium  Ethacrynic acid  Furosemide  Torsemide  Amiloride  Triamterene |
| RAAS Inhibitors | Benazepril  Captopril  Enalapril  Fosinopril  Lisinopril  Moexipril  Perindopril  Quinapril  Ramipril  Trandolapril  Azilsartan  Candesartan  Eprosartan  Irbesartan  Losartan  Olmesartan  Telmisartan  Valsartan  Aliskiren |
| Beta Blockers | Atenolol  Betaxolol  Bisoprolol  Metoprolol tartrate  Metoprolol succinate  Nebivolol  Nadolol  Propanolol  Acebutolol  Pindolol  Timolol  Carvedilol  Labetalol  Esmolol  Sotalol |
| Calcium Blockers | Amlodipine  Felodipine  Isradipine  Nicardipine  Nifedipine  Nisoldipine  Clevidipine  Nimodipine  Diltiazem  Verapamil |
| Other antihypertensive medications | Doxazosin  Prazosin  Terazosin  Alfuzosin  Clonidine  Methyldopa  Guanfacine  Hydralazine  Minoxidil  Guenethidine  Tolazoline  Sodium nitroprusside  Phenoxybenzamine hydrochloride  Phentolamine  Fenoldopam |
| NSAID, **Nonsteroidal Anti-Inflammatory Drug** | Ibuprofen  Naproxen  Aspirin  Celecoxib  Diclofenac  Indomethacin  Ketorolac  Meloxicam  Piroxicam  Etodolac  Nabumetone  Oxaprozin  Sulindac  Ketoprofen  Mefenamic acid  Flurbiprofen  Fenoprofen  Tolmetin |

**Supplementary Table S4.** Cohort Selection Process, Inclusion and Exclusion Criteria, and Final Analytic Sample for VINT and Usual Care Cohorts

| **Inclusion Criteria for Analysis** | **VINT** | **UC** | **Exclusion and Data Availability** |
| --- | --- | --- | --- |
| **Initial Population Available** | **115,942** | **2,390,947** |  |
|  | **N= 101,566** | **N=1,663,917** | **Did not meet closed claims requirement** |
| **Met closed claims requirement with ≥1 year pre-enrollment/index and continuous claims criteria (< 30 days gap)** | **14,376** | **727,030** |  |
|  | N=0 | N=0 | ESRD/dialysis |
|  | N=104 | N=15,598 | Active or metastatic cancer |
|  | N=33 | N=3912 | Transplant status |
|  | N=163 | N=46,348 | Hospice or critical care |
|  | N=77 | N=10,690 | Neurodegenerative disease |
|  | N=33 | N=1,584 | Respiratory support |
|  | N=1 | N=125 | Advanced heart failure and support |
|  | N=101 | N=17,287 | Advanced liver disease exclusion |
|  | **N= 424** | **N= 68,291** | **Comprehensive all of them together for exclusion** |
| **Cohort after excluding severe comorbid conditions** | **13,952** | **658,739** |  |
|  | **N=85** | **N=8.823** | **Type 1 diabetes diagnoses** |
| **Cohort after excluding Type 1 diabetes** | **13,867** | **649,916** |  |
|  | **N=8** | **N=31,881** | **Age at enrollment or index date < 18 years** |
| **Cohort with age at enrollment or index date 18 year or greater** | **13,859** | **618,035** |  |
|  | **N=1,969** | **N= 169,032** | **Claims follow-up less than 6 months** |
| **Cohort with at least of 6 months of claims follow-up** | **11,890** | **310,889** |  |
|  | **N=193** | **N=7,451** | **Other kidney diseases** |
| **Cohort after excluding other kidney diseases** | **11,697** | **303,438** |  |
|  | **N= 9** | **N=0** | **Removed duplicates PATIENT ID** |
|  | **11,688** | **303,438** |  |
|  | **N= 463** | **N= 29,983** | **Removed baseline chronic kidney diseases and albuminuria** |
| **Final cohort for propensity score matching and IPTW** | **11,225** | **274,455** |  |
| **Propensity Score Matching (Tier 3 – Most Stringent)**  **1:1 nearest neighbor matching (no replacement)**  **Based on logit of propensity score**  **Exact matching on:**  **Sex**  **Age category**  **Race/ethnicity**  **Diabetes**  **Obesity**  **Kidney stones**  **Index year**  **Covariates included:**  **Clinical comorbidities including CVD, CAD, Hyperlipidemia, Hypertension, etc**  **Medication use**  **Healthcare utilization & costs**  **Census region and ADI** | | | |
| **Final propensity-score matched cohort** | **11,077** | **11,077** |  |

**Supplementary Table S5.** Demographics and treatment characteristics of matched VINT participants used in this analysis versus unmatched VINT participants or those without closed claims.

| Variables | Excluded Cohort, n=110,288 | Matched Final Analytic Cohort, n=11,077 | SMD |
| --- | --- | --- | --- |
|  | **Mean (SD) or n (%)** | **Mean (SD) or n (%)** |  |
| Age (years) | 50.7 (11.0) | 50.9 (10.23) | 0.02 |
| Gender |  |  | 0.06 |
| Female | 65778 (62.7) | 6655 (60.1) |  |
| Male | 38502 (36.7) | 4372 (39.5) |  |
| Unknown | 628 (0.6) | 53 (0.5) |  |
| Race/Ethnicity |  |  | 0.23 |
| White | 39344 (51.9) | 5594 (61.6) |  |
| African American | 9944 (13.1) | 1192 (13.1) |  |
| Hispanic or Latino | 3243 (4.3) | 349 (3.8) |  |
| Asian or Pacific Islander | 10547 (13.9) | 923 (10.2) |  |
| Other | 2684 (3.5) | 228 (2.5) |  |
| Unknown | 10089 (13.3) | 790 (8.7) |  |
| Enrollment BMI Category (%) |  |  | 0.12 |
| <18.5 | 53 (0.0) | 1 (0.0) |  |
| 18.5-24.9 | 4017 (3.7) | 294 (2.7) |  |
| 25.0-29.9 | 20137 (18.4) | 1650 (14.9) |  |
| 30.0-34.9 | 34790 (31.8) | 3773 (34.1) |  |
| 35.0-39.9 | 25103 (22.9) | 2698 (24.4) |  |
| ≥40 | 25310 (23.1) | 2651 (24.0) |  |
| Enrollment HbA1c Category (%) |  |  | 0.10 |
| <6.5% | 52957 (60.8) | 5767 (65.2) |  |
| 6.5-<7.0% | 9259 (10.6) | 889 (10.0) |  |
| 7.0-<8.0% | 11187 (12.8) | 1001 (11.3) |  |
| 8.0-<9.0% | 5963 (6.8) | 515 (5.8) |  |
| ≥9% | 7805 (9.0) | 679 (7.7) |  |
| Tenure in Virta Program (%) |  |  | 0.10 |
| < 6 months | 17981 (16.3) | 1603 (14.5) |  |
| 6 to 12 months | 24117 (21.9) | 2077 (18.7) |  |
| 12 to 24 months | 42245 (38.3) | 4573 (41.3) |  |
| ≥ 24 months | 25945 (23.5) | 2827 (25.5) |  |
| Current Virta Program |  |  | 0.28 |
| Diabetes Reversal | 45686 (42.0) | 4067 (36.9) |  |
| Sustainable Weight Loss | 63,079 (57.2) | 6,951 (62.8) |  |
| 6 Months Mean BHB Category |  |  | 0.08 |
| 0-0.3mM | 26396 (31.5) | 2588 (28.0) |  |
| 0.3-0.5mM | 24592 (29.3) | 2824 (30.5) |  |
| 0.5-1.0mM | 25305 (30.2) | 2924 (31.6) |  |
| ≥ 1mM | 7628 (9.1) | 908 (9.8) |  |
| 6 Months Percentage Weight Change Category |  |  | 0.06 |
| <5% | 21991 (36.5) | 2423 (34.0) |  |
| 5-<10% | 16367 (27.2) | 1942 (27.3) |  |
| 10-<15% | 12284 (20.4) | 1508 (21.2) |  |
| 15-<20% | 6244 (10.4) | 814 (11.4) |  |
| ≥20% | 3340 (5.5) | 437 (6.1) |  |
| 12 Months Percentage Weight Change Category |  |  | 0.06 |
| <5% | 13939 (39.5) | 1468 (37.2) |  |
| 5-<10% | 8396 (23.8) | 942 (23.9) |  |
| 10-<15% | 5859 (16.6) | 652 (16.5) |  |
| 15-<20% | 3622 (10.3) | 439 (11.1) |  |
| ≥20% | 3445 (9.8) | 443 (11.2) |  |
| 6 Months HbA1c Change Category (%) |  |  | 0.07 |
| <0.5% Decrease | 18155 (49.3) | 2164 (52.6) |  |
| 0.5-1.0% Decrease | 7239 (19.7) | 800 (19.4) |  |
| ≥1.0% Decrease | 11396 (31.0) | 1153 (28.0) |  |
| 12 Months HbA1c Change Category (%) |  |  | 0.05 |
| <0.5% Decrease | 11884 (52.9) | 1405 (55.0) |  |
| 0.5-1.0% Decrease | 4050 (18.0) | 461 (18.1) |  |
| ≥1.0% Decrease | 6515 (29.0) | 687 (26.9) |  |
| 6 months diabetes reversal (%) | 22160 (56.2) | 2866 (64.5) | 0.17 |
| 12 months diabetes reversal (%) | 12429 (52.8) | 1663 (61.7) | 0.18 |
| 6 months diabetes remission (%) | 15173 (38.5) | 2007 (45.2) | 0.14 |
| 12 months diabetes remission (%) | 8581 (36.4) | 1192 (44.2) | 0.16 |
| 24 months diabetes remission (%) | 1789 (30.3) | 195 (31.6) | 0.03 |
| Index Year |  |  | 0.29 |
| 2015 | 234 (0.2) | 4 (0.0) |  |
| 2016 | 223 (0.2) | 3 (0.0) |  |
| 2017 | 621 (0.6) | 12 (0.1) |  |
| 2018 | 1020 (0.9) | 18 (0.2) |  |
| 2019 | 1883 (1.7) | 186 (1.7) |  |
| 2020 | 4013 (3.7) | 498 (4.5) |  |
| 2021 | 10206 (9.5) | 793 (7.2) |  |
| 2022 | 14573 (13.5) | 1731 (15.6) |  |
| 2023 | 15963 (14.8) | 1829 (16.5) |  |
| 2024 | 42830 (39.8) | 5091 (45.9) |  |
| 2025 | 16092 (14.9) | 915 (8.3) |  |

**Note:** Standardized mean differences (SMDs) and *p*-values were calculated using the TableOne package in R.To evaluate covariate balance between matched groups, SMD ≤ 0.1 was used as the criterion for acceptable balance.

**Abbreviations:** UC, usual care; VINT, Individualized Nutrition Therapy program; BMI, Body mass index; HbA1c, hemoglobin A1c

**Supplementary Table S6.** Baseline characteristics of unmatched and propensity score matched secondary cohorts of VINT (with at least 6m VINT tenure) and UC

| Variables | Unmatched Dataset | | | Matched Dataset | | |
| --- | --- | --- | --- | --- | --- | --- |
|  | **VINT (n=11,225)** | **UC (n=274,455)** | **SMD** | **VINT (n=9.740)** | **UC (n=9,740)** | **SMD** |
|  | **Mean (SD)**  **or n (%)** | **Mean (SD)**  **or n (%)** |  | **Mean (SD)**  **or n (%)** | **Mean (SD)**  **or n (%)** |  |
| Demographics and Characteristics | | | | | | |
| Age (years) | 50.7 (10.2) | 53.3 (16.0) | 0.20 | 51.6 (10.1) | 51.5 (9.9) | 0.01 |
| Follow duration (days) | 686.6 (397.0) | 576.65 (410.2) | 0.27 | 680.0 (417.8) | 721.9 (415.1) | 0.10 |
| Gender |  |  | 0.04 |  |  | <0.001 |
| Female | 6722 (59.9) | 160582 (58.5) |  | 5788 (59.4) | 5788 (59.4) |  |
| Male | 4433 (39.5) | 111443 (40.6) |  | 3910 (40.1) | 3910 (40.1) |  |
| Unknown | 70 (0.6) | 2430 (0.9) |  | 42 (0.4) | 42 (0.4) |  |
| Race and Ethnicity |  |  | 0.14 |  |  | 0.09 |
| White | 5657 (61.5) | 134755 (57.5) |  | 4598 (59.5) | 5009 (62.3) |  |
| African American | 1214 (13.2) | 34461 (14.7) |  | 992 (12.8) | 1064 (13.2) |  |
| Hispanic or Latino | 938 (10.2) | 30947 (13.2) |  | 753 (9.7) | 795 (9.9) |  |
| Asian or Pacific Islander | 351 (3.8) | 8436 (3.6) |  | 342 (4.4) | 315 (3.9) |  |
| Other | 235 (2.6) | 8386 (3.6) |  | 255 (3.3) | 196 (2.4) |  |
| Unknown | 803 (8.7) | 17193 (7.3) |  | 794 (10.3) | 666 (8.3) |  |
| Baseline Comorbidities | | | | | | |
| Type 2 diabetes | 4086 (36.4) | 63283 (23.1) | 0.30 | 3992 (41.0) | 3992 (41.0) | <0.001 |
| Obesity | 6135 (54.7) | 125775 (45.8) | 0.18 | 5183 (53.2) | 5183 (53.2) | <0.001 |
| Chronic Kidney Disease | 0 (0.0) | 0 (0.0) | <0.001 | 0 (0.0) | 0 (0.0) | <0.001 |
| Kidney Stones | 179 (1.6) | 5525 (2.0) | 0.03 | 9740(100.0) | 9740 (100.0) | <0.001 |
| Heart Failure | 87 (0.8) | 8874 (3.2) | 0.18 | 0.00 (0.00) | 0.00 (0.00) | <0.001 |
| Hyperlipidemia | 4957 (44.2) | 126180 (46.0) | 0.04 | 112 (1.1) | 112 (1.1) | <0.001 |
| Hypertension | 4757 (42.4) | 126826 (46.2) | 0.08 | 82 (0.8) | 85 (0.9) | 0.003 |
| Stroke | 153 (1.4) | 10059 (3.7) | 0.15 | 4547 (46.7) | 4552 (46.7) | 0.001 |
| Coronary artery disease | 484 (4.3) | 23203 (8.5) | 0.17 | 4365 (44.8) | 4383 (45.0) | 0.004 |
| Cardiovascular disease | 66 (0.6) | 3968 (1.4) | 0.09 | 136 (1.4) | 140 (1.4) | 0.003 |
| Smoking | 364 (3.2) | 17622 (6.4) | 0.15 | 398 (4.1) | 442 (4.5) | 0.02 |
| Baseline Medications | | | | | | |
| SGLT2i | 908 (8.1) | 11301 (4.1) | 0.17 | 877 (9.0) | 877 (9.0) | <0.001 |
| Incretin Mimetics | 1993 (17.8) | 31395 (11.4) | 0.18 | 1931 (19.8) | 1931 (19.8) | <0.001 |
| RAASi | 3701 (33.0) | 92801 (33.8) | 0.02 | 3743 (38.4) | 3743 (38.4) | <0.001 |
| MRA and nonMRA | 289 (2.6) | 7960 (2.9) | 0.02 | 238 (2.4) | 238 (2.4) | <0.001 |
| Diuretic | 2014 (17.9) | 60152 (21.9) | 0.10 | 1995 (20.5) | 2004 (20.6) | 0.002 |
| Beta blocker | 1224 (10.9) | 47955 (17.5) | 0.19 | 1246 (12.8) | 1243 (12.8) | 0.001 |
| Calcium blocker | 1282 (11.4) | 43893 (16.0) | 0.13 | 1216 (12.5) | 1285 (13.2) | 0.021 |
| Other antihypertensive medications | 230 (2.0) | 9051 (3.3) | 0.08 | 213 (2.2) | 228 (2.3) | 0.01 |
| Insulin | 605 (5.4) | 10047 (3.7) | 0.08 | 645 (6.6) | 620 (6.4) | 0.01 |
| Sulfonylureas | 339 (3.0) | 4901 (1.8) | 0.08 | 351 (3.6) | 344 (3.5) | 0.004 |
| DPP4i | 384 (3.4) | 5022 (1.8) | 0.10 | 434 (4.5) | 378 (3.9) | 0.03 |
| Thiazolidine | 195 (1.7) | 2942 (1.1) | 0.06 | 190 (2.0) | 195 (2.0) | 0.004 |
| Other T2D medications | 3266 (29.1) | 46767 (17.0) | 0.29 | 3330 (34.2) | 3287 (33.7) | 0.01 |
| Baseline cost and trajectory | | | | | | |
| Total cost of care | 11773.0 (24464.4) | 10649.2 (22926.6) | 0.05 | 11815.8 (21345.9) | 12073.8 (24686.4) | 0.01 |
| Outpatient cost | 904.5 (7410.2) | 1157.1 (7761.8) | 0.03 | 897.6 (5876.3) | 899.9 (7388.9) | <0.001 |
| Inpatient cost | 3638.8 (12464.6) | 3214.3 (12949.7) | 0.03 | 3978.09 (9932.1) | 3832.7 (13108.5) | 0.01 |
| Pharmacy cost | 7227.6 (17690.8) | 6271.3 (14965.2) | 0.06 | 6939.31 (16094.5) | 7338.66 (17595.3) | 0.02 |
| Baseline area level variables | | | | | | |
| Region |  |  | 0.48 |  |  | 0.045 |
| Midwest | 4191 (37.3) | 73652 (26.8) |  | 3582 (36.8) | 3672 (37.7) |  |
| Northeast | 999 (8.9) | 70130 (25.6) |  | 1004 (10.3) | 876 (9.0) |  |
| South | 4680 (41.7) | 91298 (33.3) |  | 3996 (41.0) | 4023 (41.3) |  |
| West | 1355 (12.1) | 39371 (14.3) |  | 1158 (11.9) | 1169 (12.0) |  |
| Area Deprivation Index (ADI) Quintiles | | | 0.29 |  |  | 0.04 |
| ADI 1 | 1840 (16.4) | 70001 (25.5) |  | 1703 (17.5) | 1611 (16.5) |  |
| ADI 2 | 4704 (41.9) | 85152 (31.0) |  | 3883 (39.9) | 4056 (41.6) |  |
| ADI 3 | 2832 (25.2) | 64178 (23.4) |  | 2507 (25.7) | 2496 (25.6) |  |
| ADI 4 | 1168 (10.4) | 34275 (12.5) |  | 1024 (10.5) | 995 (10.2) |  |
| ADI 5 | 681 (6.1) | 20844 (7.6) |  | 623 (6.4) | 582 (6.0) |  |
| Index Year |  |  | 0.56 |  |  | <0.001 |
| 2011 | 0 (0.0) | 383 (0.1) |  | 0 (0.0) | 0 (0.0) |  |
| 2012 | 0 (0.0) | 490 (0.2) |  | 0 (0.0) | 0 (0.0) |  |
| 2013 | 0 (0.0) | 1550 (0.6) |  | 0 (0.0) | 0 (0.0) |  |
| 2014 | 0 (0.0) | 832 (0.3) |  | 0 (0.0) | 0 (0.0) |  |
| 2015 | 3 (0.0) | 1484 (0.5) |  | 10 (0.1) | 10 (0.1) |  |
| 2016 | 3 (0.0) | 2601 (0.9) |  | 3 (0.0) | 3 (0.0) |  |
| 2017 | 12 (0.1) | 6252 (2.3) |  | 12 (0.1) | 12 (0.1) |  |
| 2018 | 19 (0.2) | 5144 (1.9) |  | 19 (0.2) | 19 (0.2) |  |
| 2019 | 201 (1.8) | 13814 (5.0) |  | 198 (2.0) | 198 (2.0) |  |
| 2020 | 521 (4.6) | 19707 (7.2) |  | 517 (5.3) | 517 (5.3) |  |
| 2021 | 812 (7.2) | 28003 (10.2) |  | 822 (8.4) | 822 (8.4) |  |
| 2022 | 1768 (15.8) | 41983 (15.3) |  | 1793 (18.4) | 1793 (18.4) |  |
| 2023 | 1856 (16.5) | 41482 (15.1) |  | 1851 (19.0) | 1851 (19.0) |  |
| 2024 | 5108 (45.5) | 71585 (26.1) |  | 3800 (39.0) | 3800 (39.0) |  |
| 2025 | 922 (8.2) | 39145 (14.3) |  | 715 (7.3) | 715 (7.3) |  |
| Variables not included in the matching | | | | | | |
| Other baseline comorbidities | | | | | | |
| Cancer | 446 (4.0) | 15953 (5.8) | 0.09 | 442 (4.5) | 412 (4.2) | 0.02 |
| Afib | 172 (1.5) | 11163 (4.1) | 0.15 | 171 (1.8) | 159 (1.6) | 0.01 |
| Glaucoma | 444 (4.0) | 16377 (6.0) | 0.09 | 401 (4.1) | 412 (4.2) | 0.01 |
| MASLD | 655 (5.8) | 14058 (5.1) | 0.03 | 573 (5.9) | 572 (5.9) | <0.001 |
| MASH | 55 (0.5) | 1385 (0.5) | 0.002 | 63 (0.6) | 45 (0.5) | 0.03 |
| Liver fibrosis | 25 (0.2) | 419 (0.2) | 0.02 | 15 (0.2) | 21 (0.2) | 0.01 |
| GERD | 1566 (14.0) | 48361 (17.6) | 0.10 | 1681 (17.3) | 1382 (14.2) | 0.08 |
| Osteoarthritis | 1427 (12.7) | 48113 (17.5) | 0.14 | 1464 (15.0) | 1286 (13.2) | 0.05 |
| Sleep Disorder | 2751 (24.5) | 56551 (20.6) | 0.09 | 2193 (22.5) | 2455 (25.2) | 0.06 |
| PAD | 167 (1.5) | 14450 (5.3) | 0.21 | 267 (2.7) | 166 (1.7) | 0.07 |
| Diabetes Neuropathy | 387 (3.4) | 10895 (4.0) | 0.03 | 501 (5.1) | 404 (4.1) | 0.05 |
| Schizophrenia | 18 (0.2) | 2307 (0.8) | 0.10 | 71 (0.7) | 18 (0.2) | 0.08 |
| Bipolar disorder | 127 (1.1) | 5544 (2.0) | 0.07 | 210 (2.2) | 100 (1.0) | 0.09 |
| Common mental health | 2490 (22.2) | 62832 (22.9) | 0.02 | 2404 (24.7) | 2063 (21.2) | 0.08 |
| Other mental health conditions | 814 (7.3) | 17270 (6.3) | 0.04 | 651 (6.7) | 659 (6.8) | 0.003 |
| Other baseline medications | | | | | | |
| Anticoagulant | 227 (2.0) | 13181 (4.8) | 0.15 | 294 (3.0) | 232 (2.4) | 0.04 |
| Antiplatelet | 291 (2.6) | 13536 (4.9) | 0.12 | 391 (4.0) | 298 (3.1) | 0.05 |
| Statin | 3577 (31.9) | 91963 (33.5) | 0.04 | 3587 (36.8) | 3591 (36.9) | 0.001 |
| PCSK9i | 531 (4.7) | 13553 (4.9) | 0.01 | 36 (0.4) | 36 (0.4) | <0.001 |
| Other lipid lowering medications | 37 (0.3) | 1152 (0.4) | 0.02 | 580 (6.0) | 532 (5.5) | 0.02 |
| Sacubitril | 15 (0.1) | 1241 (0.5) | 0.06 | 22 (0.2) | 14 (0.1) | 0.02 |
| Neuropathy medications | 1548 (13.8) | 51526 (18.8) | 0.14 | 1964 (20.2) | 1542 (15.8) | 0.11 |
| Antidepressant | 2944 (26.2) | 76229 (27.8) | 0.04 | 2962 (30.4) | 2883 (29.6) | 0.02 |
| Antipsychotic | 197 (1.8) | 10793 (3.9) | 0.13 | 394 (4.0) | 191 (2.0) | 0.12 |

**Note:** Standardized mean differences (SMDs) were computed as the absolute difference in means (or proportions for categorical variables) between groups divided by the pooled standard deviation. SMDs were calculated using both the TableOne and MatchIt packages to cross-validate covariate balance estimates. To evaluate covariate balance between matched groups, SMD ≤ 0.1 was used as the criterion for acceptable balance. Unlike *p*-values, which are influenced by sample size and may indicate statistically significant differences that are not clinically meaningful in large samples, SMD provides a sample size–independent measure of balance that quantifies the magnitude of difference between groups. Therefore, SMDs were used as the primary metric for assessing post-matching balance, while *p*-values were reported descriptively.

**Abbreviations:** UC, usual care; VINT, Individualized Nutrition Therapy (intervention) program; T2D, type 2 diabetes; MASLD, metabolic dysfunction-associated steatotic liver disease; MASH, metabolic dysfunction-associated steatohepatitis; GERD, gastro-oesophageal reflux disease; Afib, atrial fibrillation; PAD, peripheral artery disease; CKD, chronic kidney disease; SGLT2i, sodium–glucose cotransporter-2 inhibitor; DPP4i, dipeptidyl peptidase-4 inhibitor; RAASi, renin–angiotensin–aldosterone system inhibitor (e.g., ACEi/ARB/ARNI); MRA, mineralocorticoid receptor antagonist (steroidal); non-MRA, non-steroidal mineralocorticoid receptor antagonist (e.g., finerenone); PCSK9i, proprotein convertase subtilisin/kexin type 9 inhibitor.

**Supplementary Table S7.** Cox regression models for all new-onset of chronic kidney disease (CKD) and stage 3 CKD and beyond, presenting the effect estimates after covariate adjustment in the primary 1:1 matched cohort analysis of VINT and UC

| Variables | Factor | All new-onset CKD | | | Stage 3 CKD and beyond | | |
| --- | --- | --- | --- | --- | --- | --- | --- |
|  |  | **aHR** | **95% CI** | **p-value** | **aHR** | **95% CI** | **p-value** |
| Treatment | **UC** | Referent |  |  | Referent |  |  |
|  | **VINT** | 0.68 | (0.56–0.82) | <0.001 | 0.60 | (0.47-0.77) | <0.001 |
| Age at registration |  | 1.07 | (1.06–1.08) | <0.001 | 1.09 | (1.07-1.11) | <0.001 |
| Gender | **Male** | Referent |  |  | Referent |  |  |
|  | **Female** | 0.83 | (0.69–1.01) | 0.06 | 0.82 | (0.64-1.05) | 0.11 |
| Race/Ethnicity | **White** | Referent |  |  | Referent |  |  |
|  | **Asian/Pacific Islander** | 1.19 | (0.72–1.98) | 0.50 | 0.94 | (0.44-2.02) | 0.88 |
|  | **Black/African American** | 1.32 | (1.03–1.70) | 0.03 | 1.44 | (1.05-1.96) | 0.02 |
|  | **Hispanic/Latino** | 1.39 | (1.01–1.90) | 0.04 | 1.29 | (0.84-1.97) | 0.24 |
|  | **Other** | 1.49 | (0.88–2.51) | 0.14 | 1.37 | (0.67-2.80) | 0.39 |
|  | **Unknown** | 0.98 | (0.66–1.47) | 0.93 | 1.04 | (0.62-1.75) | 0.88 |
| T2D | **No** | Referent |  |  | Referent |  |  |
|  | **Yes** | 1.35 | (1.05–1.74) | 0.02 | 1.13 | (0.82-1.57) | 0.46 |
| Obesity | **No** | Referent |  |  | Referent |  |  |
|  | **Yes** | 1.08 | (0.89–1.31) | 0.42 | 1.09 | (0.85-1.39) | 0.50 |
| Medication adherence (PDC) | **MRA/non-MRA** | 2.42 | (1.64–3.56) | <0.001 | 2.85 | (1.81-4.49) | <0.001 |
|  | **GLP-1RA** | 1.14 | (0.89–1.47) | 0.30 | 1.38 | (1.00-1.91) | 0.05 |
|  | **SGLT2i** | 2.10 | (1.61–2.73) | <0.001 | 2.16 | (1.55-3.02) | <0.001 |
|  | **RAASi** | 1.33 | (1.03–1.72) | 0.03 | 1.59 | (1.13-2.24) | 0.01 |
|  | **Diuretic** | 2.06 | (1.64–2.59) | <0.001 | 2.37 | (1.77-3.17) | <0.001 |
|  | **NSAID** | 2.40 | (1.84–3.12) | <0.001 | 2.16 | (1.54-3.04) | <0.001 |

**Note:** Cox proportional hazards regression models were used to estimate hazard ratios (HRs) and 95% confidence intervals (CIs) for the association between treatment group (VINT vs. UC) and risk of new-onset chronic kidney disease (CKD) and albuminuria as well as stage 3 CKD and beyond. The matched analysis was based on the primary 1:1 propensity score–matched cohort. All models were adjusted for age (per 1-year increase), sex, race/ethnicity, type 2 diabetes, obesity, SGLT2 inhibitor use, GLP-1 receptor agonist use, RAAS inhibitor use, mineralocorticoid receptor antagonist (MRA/non-MRA) use, diuretic use, and NSAID use. White participants were used as the reference race/ethnicity category. HRs >1 indicate increased risk and HRs <1 indicate reduced risk relative to the reference group. p-values <0.05 were considered statistically significant. **Abbreviations:** UC, usual care; VINT, Individualized Nutrition Therapy (intervention) program; HR, hazard ratio; CI, confidence interval; CKD, chronic kidney disease; T2D, type 2 diabetes; ACEI, angiotensin-converting enzyme inhibitor; ARB, angiotensin II receptor blocker; ACEI/ARB, angiotensin-converting enzyme inhibitor or angiotensin II receptor blocker; SGLT2i, sodium–glucose cotransporter-2 inhibitor; MRA, mineralocorticoid receptor antagonist; non-MRA, non-steroidal mineralocorticoid receptor antagonist (e.g., finerenone); BMI, body mass index, NSAID, **Nonsteroidal Anti-Inflammatory Drug**

**Supplementary Table S8.** Cox regression models for all new-onset of chronic kidney disease (CKD) and stage 3 CKD and beyond, presenting the effect estimates after covariate adjustment in the secondary 1:1 matched cohort analysis of VINT and UC where VINT participants with ≥6 months of program engagement

| Variables | Factor | All new-onset CKD | | | Stage 3 CKD and beyond | | |
| --- | --- | --- | --- | --- | --- | --- | --- |
|  |  | **aHR** | **95% CI** | **p-value** | **aHR** | **95% CI** | **p-value** |
| Treatment | **UC** | Referent |  |  | Referent |  |  |
|  | **VINT** | 0.61 | (0.50–0.74) | <0.001 | 0.52 | (0.40–0.68) | <0.001 |
| Age at registration |  | 1.07 | (1.06–1.08) | <0.001 | 1.09 | (1.07–1.11) | <0.001 |
| Gender | **Male** | Referent |  |  | Referent |  |  |
|  | **Female** | 0.81 | (0.67–0.99) | 0.04 | 0.84 | (0.65–1.08) | 0.17 |
| Race/Ethnicity | **White** | Referent |  |  | Referent |  |  |
|  | **Asian/Pacific Islander** | 0.88 | (0.51–1.51) | 0.64 | 1.00 | (0.47–2.12) | 1.00 |
|  | **Black/African American** | 0.81 | (0.46–1.45) | 0.48 | 0.87 | (0.39–1.94) | 0.74 |
|  | **Hispanic/Latino** | 0.73 | (0.35–1.49) | 0.38 | 0.59 | (0.20–1.71) | 0.33 |
|  | **Other** | 0.60 | (0.32–1.12) | 0.11 | 0.54 | (0.22–1.32) | 0.18 |
|  | **Unknown** | 0.65 | (0.34–0.94) | 0.03 | 0.60 | (0.29–1.23) | 0.16 |
| T2D | **No** | Referent |  |  | Referent |  |  |
|  | **Yes** | 0.74 | (0.57–0.96) | 0.03 | 0.63 | (0.44–0.89) | 0.01 |
| Obesity | **No** | Referent |  |  | Referent |  |  |
|  | **Yes** | 1.19 | (0.98–1.44) | 0.08 | 1.20 | (0.94–1.55) | 0.15 |
| Medication adherence (PDC) | **MRA/non-MRA** | 2.68 | (1.80–3.98) | <0.001 | 3.22 | (2.03–5.12) | <0.001 |
|  | **GLP-1RA** | 1.52 | (1.17–1.99) | 0.002 | 1.68 | (1.19–2.37) | 0.003 |
|  | **SGLT2i** | 1.93 | (1.48–2.53) | <0.001 | 2.32 | (1.65–3.27) | <0.001 |
|  | **RAASi** | 1.23 | (0.94–1.61) | 0.13 | 1.37 | (0.95–1.97) | 0.09 |
|  | **Diuretic** | 1.81 | (1.43–2.29) | <0.001 | 2.23 | (1.64–3.02) | <0.001 |
|  | **NSAID** | 2.52 | (1.93–3.29) | <0.001 | 2.01 | (1.42–2.85) | <0.001 |

**Note:** Cox proportional hazards regression models were used to estimate hazard ratios (HRs) and 95% confidence intervals (CIs) for the association between treatment group (VINT vs. UC) and risk of new-onset chronic kidney disease (CKD) and albuminuria as well as stage 3 CKD and beyond. The matched analysis was based on the secondary 1:1 propensity score–matched cohort. All models were adjusted for age (per 1-year increase), sex, race/ethnicity, type 2 diabetes, obesity, SGLT2 inhibitor use, GLP-1 receptor agonist use, RAAS inhibitor use, mineralocorticoid receptor antagonist (MRA/non-MRA) use, diuretic use, and NSAID use. White participants were used as the reference race/ethnicity category. HRs >1 indicate increased risk and HRs <1 indicate reduced risk relative to the reference group. p-values <0.05 were considered statistically significant. **Abbreviations:** UC, usual care; VINT, Individualized Nutrition Therapy (intervention) program; HR, hazard ratio; CI, confidence interval; CKD, chronic kidney disease; T2D, type 2 diabetes; ACEI, angiotensin-converting enzyme inhibitor; ARB, angiotensin II receptor blocker; ACEI/ARB, angiotensin-converting enzyme inhibitor or angiotensin II receptor blocker; SGLT2i, sodium–glucose cotransporter-2 inhibitor; MRA, mineralocorticoid receptor antagonist; non-MRA, non-steroidal mineralocorticoid receptor antagonist (e.g., finerenone); BMI, body mass index, NSAID, **Nonsteroidal Anti-Inflammatory Drug**

**Supplementary Table S9.** Cox regression models for all new-onset of chronic kidney disease (CKD) and stage 3 CKD and beyond, presenting the effect estimates after covariate adjustment in VINT and UC sensitivity analyses (IPTW-weighted model)

| Variables | Factor | All new-onset CKD | | | Stage 3 CKD and beyond | | |
| --- | --- | --- | --- | --- | --- | --- | --- |
|  |  | **aHR** | **95% CI** | **p-value** | **aHR** | **95% CI** | **p-value** |
| Treatment | **UC** | Referent |  |  | Referent |  |  |
|  | **VINT** | 0.59 | (0.48–0.73) | <0.001 | 0.56 | (0.43-0.73) | <0.001 |
| Age at registration |  | 1.08 | (1.07–1.08) | <0.001 | 1.09 | (1.08-1.09) | <0.001 |
| Gender | **Male** | Referent |  |  | Referent |  |  |
|  | **Female** | 0.93 | (0.90–0.97) | 0.001 | 1.00 | (0.95-1.05) | 0.954 |
|  | **Unknown** | 0.99 | (0.78–1.26) | 0.911 | 0.88 | (0.64-1.20) | 0.423 |
| Race/Ethnicity | **White** | Referent |  |  | Referent |  |  |
|  | **Asian/Pacific Islander** | 0.90 | (0.78–1.04) | 0.152 | 0.80 | (0.65-0.98) | 0.029 |
|  | **Black/African American** | 1.47 | (1.40–1.55) | <0.001 | 1.39 | (1.31-1.49) | <0.001 |
|  | **Hispanic/Latino** | 1.09 | (1.01–1.17) | 0.019 | 0.89 | (0.82-0.98) | 0.018 |
|  | **Other** | 1.05 | (0.92–1.20) | 0.453 | 1.07 | (0.91-1.27) | 0.416 |
|  | **Unknown** | 0.78 | (0.71–0.85) | <0.001 | 0.82 | (0.74-0.92) | <0.001 |
| T2D | **No** | Referent |  |  | Referent |  |  |
|  | **Yes** | 1.13 | (1.08–1.18) | <0.001 | 1.09 | (1.03-1.16) | 0.003 |
| Obesity | **No** | Referent |  |  | Referent |  |  |
|  | **Yes** | 1.08 | (1.04–1.13) | <0.001 | 0.98 | (0.93-1.03) | 0.384 |
| Medication adherence (PDC) | **MRA/non-MRA** | 2.28 | (2.11–2.47) | <0.001 | 2.58 | (2.36-2.83) | <0.001 |
|  | **GLP-1RA** | 1.34 | (1.25–1.44) | <0.001 | 1.39 | (1.28-1.51) | <0.001 |
|  | **SGLT2i** | 2.05 | (1.91–2.20) | <0.001 | 2.23 | (2.05-2.43) | <0.001 |
|  | **RAASi** | 1.55 | (1.47–1.64) | <0.001 | 1.58 | (1.48-1.69) | <0.001 |
|  | **Diuretic** | 1.72 | (1.64–1.81) | <0.001 | 1.97 | (1.86-2.10) | <0.001 |
|  | **NSAID** | 2.14 | (2.02–2.27) | <0.001 | 2.04 | (1.89-2.19) | <0.001 |

**Note:** Cox proportional hazards regression models were used to estimate hazard ratios (HRs) and 95% confidence intervals (CIs) for the association between treatment group (VINT vs. UC) and risk of new-onset chronic kidney disease (CKD) and albuminuria. The IPTW analysis included all eligible participants and used stabilized inverse probability of treatment weighting based on the same demographic, clinical, medication, healthcare utilization, and socioeconomic covariates used in the propensity score matching model. All models were adjusted for age (per 1-year increase), sex, race/ethnicity, type 2 diabetes, obesity, SGLT2 inhibitor use, GLP-1 receptor agonist use, RAAS inhibitor use, mineralocorticoid receptor antagonist (MRA/non-MRA) use, diuretic use, and NSAID use. White participants were used as the reference race/ethnicity category. The "Unknown" sex category appears only in the IPTW analysis because the weighted cohort retained all eligible participants, including those with missing sex information. HRs >1 indicate increased risk and HRs <1 indicate reduced risk relative to the reference group. p-values <0.05 were considered statistically significant. **Abbreviations:** UC, usual care; VINT, Individualized Nutrition Therapy (intervention) program; HR, hazard ratio; CI, confidence interval; CKD, chronic kidney disease; IPTW, inverse probability of treatment weighting; T2D, type 2 diabetes; ACEI, angiotensin-converting enzyme inhibitor; ARB, angiotensin II receptor blocker; ACEI/ARB, angiotensin-converting enzyme inhibitor or angiotensin II receptor blocker; SGLT2i, sodium–glucose cotransporter-2 inhibitor; MRA, mineralocorticoid receptor antagonist; non-MRA, non-steroidal mineralocorticoid receptor antagonist (e.g., finerenone); BMI, body mass index, NSAID, **Nonsteroidal Anti-Inflammatory Drug**

**Supplementary Table S10.** Comparison of Medication Use Between VINT Participants and Matched UC Controls Among Those with CKD diagnosis at follow-up with available prescription claims follow-up before censoring

| Medication Class | VINT (n=210)  N (%) | Mean Days of Use | Median Days of Use | UC (n=303) N (%) | Mean Days of Use | Median Days of Use | p-value (Users) | p-value (Days Covered) |
| --- | --- | --- | --- | --- | --- | --- | --- | --- |
| Incretin Mimetics | 123 (58.6) | 868.0 | 686 | 125 (41.7) | 703.9 | 458 | 0.0002 | 0.05 |
| SGLT2 Inhibitors | 87 (41.4) | 635.1 | 480 | 99 (33.0) | 661.5 | 420 | 0.06 | 0.76 |
| RAAS Inhibitors | 159 (75.7) | 1217.4 | 1140 | 216 (71.3) | 1015.2 | 900 | 0.40 | 0.03 |
| Diuretics | 101 (48.1) | 858.7 | 720 | 165 (54.5) | 837.4 | 660 | 0.15 | 0.89 |
| MRA / non-MRA | 22 (10.5) | 598.6 | 495 | 28 (9.3) | 533.7 | 405 | 0.78 | 0.49 |
| Finerenone | 6 (2.9) | NA | NA | 7 (2.3) | NA | NA | 0.93 | NA |
| NSAIDs | 108 (51.4) | 282.7 | 90 | 172 (57.3) | 321.6 | 97.5 | 0.22 | 0.61 |

**Note:** This table compares the proportion and duration of medication use between VINT participants and matched controls with chronic kidney disease at baseline and/or during follow-up among those with available prescription claims prior to censoring. The proportion of medication users (*N*, %) and average number of days of use were calculated for each drug class. *p*-values for the proportion of users were derived using the chi-square test or Fisher’s exact test (for small cell counts), and *p*-values for the number of days covered were calculated using the Wilcoxon rank-sum test for non-normally distributed data.

**Abbreviations:** VINT, Individualized Nutrition Therapy (intervention) program; UC, usual care; CKD, chronic kidney disease; GLP1-RA, glucagon-like peptide-1 receptor agonist; SGLT2i, sodium–glucose cotransporter-2 inhibitor; ACEi, angiotensin-converting enzyme inhibitor; ARB, angiotensin II receptor blocker; MRA, mineralocorticoid receptor antagonist; DPP4i, dipeptidyl peptidase-4 inhibitor; TZD, thiazolidinedione; NSAID, **Nonsteroidal Anti-Inflammatory Drug**
